# Feasibility testing and preliminary trial of a crisis planning and monitoring intervention to reduce compulsory readmissions: the FINCH Study

**DOI:** 10.1101/2025.07.29.25332374

**Authors:** Sonia Johnson, Mary Birken, Rafael Gafoor, Patrick Nyikavaranda, Ariana Kular, Jordan Parkinson, Kathleen Lindsay Fraser, Jackie Hardy, Mark Keith Holden, Lizzie Mitchell, Janet Seale, Cady Stone, Valerie Christina White, Louise Blakley, Barbara Lay, Lisa Wood, Nick Freemantle, Henrietta Mbeah-Bankas, Paul McCrone, Fiona Lobban, Brynmor Lloyd-Evans

## Abstract

**Background:** Compulsory admissions to psychiatric hospital have been rising in England and some other higher income countries. Patients and families often find such admissions distressing, disempowering, and traumatising. Evidence on how to prevent compulsory admissions is still very limited. Collaborative crisis planning currently appears the most promising way of reducing compulsory readmissions, but can be challenging to implement.

**Aims:** The overall aim of the FINCH study was to develop a crisis planning and monitoring intervention to prevent repeat compulsory admissions (Phase 1) and to investigate the feasibility and acceptability of testing it through a randomised controlled trial (Phase 2).

**Methods:** Drawing on a promising intervention developed in Switzerland and on qualitative interviews with service users and carers, a team including researchers, service users, carers and clinicians co-designed an intervention. This included personalised plans for preventing and managing crises, with follow-up contacts over a year to support participants in implementing these. We carried out a feasibility randomised controlled trial of the intervention, with 80 participants recruited at the end of compulsory hospital admissions in three areas of England.

**Results:** Eighty participants were recruited within our target timeframe, 40 (as planned) from ethnic groups at disproportionately high risk of compulsory admission. Data were obtained for 86% of participants on compulsory admission, identified as a potential primary outcome for a full trial, but only for 51% on secondary outcomes measured at interview. Twenty-five of the 38 experimental group participants (66%) received at least three intervention sessions and developed a crisis plan of some kind. Qualitative data obtained from participating service users and carers suggested the intervention was acceptable and feasible, but that a high level of persistence and flexibility and considerable time were needed to deliver it.

**Conclusions:** We were readily able to recruit to this study, including from ethnic groups who are at high risk of compulsory admission, and delivery of our study intervention was feasible at least in a minimum form. Given the high financial and human costs of compulsory admission, there is an ethical and practical requirement for more research in this area: larger-scale research based on a refined version of our intervention has potential to contribute.

**Trial registration:** ISRCTN, ISRCTN11627644. Registered prospectively 25th May 2022, https://www.isrctn.com/ISRCTN11627644.

## Introduction

Compulsory detentions in mental health inpatient units have increased over several decades in England, and similar trends are observed in some other higher income countries (1). A particular concern in England is a large ethnic inequality in compulsory admissions; people from Black and Black British ethnic groups are about four times more likely to be detained compared to White British individuals, with several other minoritised ethnic groups also being at higher risk (2, 3)).

Compulsory admissions may be clinically difficult to avoid in some circumstances but are inherently coercive and should be avoided where feasible. Service users and carers frequently report that compulsory admissions are distressing and traumatising, disrupting recovery and therapeutic alliances (4–6). Human and financial burdens on clinicians and the healthcare system are also considerable with little evidence that such admissions improve longer term outcomes (7).

Evidence-based approaches to preventing compulsory admissions have yet to become a standard part of care in the UK or elsewhere. Surprisingly few trials have been conducted with compulsory admission as a primary or even secondary outcome measure (8). People who have been detained at least once are at high risk of being detained again (9), making them a promising focus for efforts to reduce compulsory detentions. One approach has been continuing compulsion into the community, such as through Community Treatment Orders in England. However, current evidence does not support this as an effective means of reducing compulsory admissions (10), and their use is disproportionately high in Black or Black British ethnic groups (11).

When evidence from all available studies internationally is pooled through meta-analysis (12, 13), the only types of intervention that currently show substantial effectiveness in reducing compulsory admissions are advance planning for crises (often called crisis plans) and collaborative agreements (advance statements) with patients about what should happen if they become unwell in the future (14,15). Such strategies were recommended for national roll-out in the Independent Review of the Mental Health Act (MHA) in England, published in 2018 (16).

In our group’s review of relevant literature (12), we found that while pooled meta-analysis indicates overall effectiveness for interventions based on crisis planning, there has been considerable variation between studies in effect size and in whether statistical significance was reached.

Challenges in implementing crisis planning strategies so that they have a sustained effect have been recurrently noted: for example in the largest UK trial, a joint crisis planning model initially effective in a single-site trial showed little evidence of effectiveness when tested across multiple sites and it was noted that crisis plans were made but rarely referred to in subsequent care or help-seeking (17–19). Thus, to be successful, crisis planning needs to be embedded in a framework that ensures plans remain salient in subsequent care, especially at the time of any future crisis.

Within our systematic review (12), we identified one study that appeared to have a more developed approach to implementation than the others, including strategies for continued monitoring for signs of crisis. This study, conducted in Zürich in Switzerland (20, 21), involved initiation of a crisis planning intervention in hospital, delivered by a “personal mental health care worker” who was a psychologist separate from the patient’s usual care team and responsible for delivering the intervention over a two year period (20). Components included three or four sessions dedicated to discussing the nature of each patient’s mental health problems and risk factors for relapse, and reinforcing their self-management skills and adherence to treatment. A crisis care plan was formulated, following which participants were monitored for two years, additional to any other mental health care provided. During this monitoring stage, personal mental health care workers contacted participants every four weeks by telephone and discussed risk factors and early warning signs for the crisis, aiming to detect an incipient relapse and to prompt action to avert it. Thus this intervention was designed to avert the often-identified problem of crisis plans being developed but subsequently forgotten(17).

Findings from implementing this Swiss model were promising: over two years, 28% of participants in this programme were compulsorily readmitted compared to 43% of patients in the control arm who received standard local care. The adjusted relative risk of compulsory readmission was 0.55 (95% confidence interval 0.33–0.94) in favour of the treatment group (20). However, differential risks of drop-out created ambiguity in interpreting the statistically significant result. Robust research is rare in this field, and the results appeared sufficiently promising to provide a starting point for our intervention. Congruent with this and other preceding studies, a further trial published after the start of the current study, and conducted in France, showed an effect in reducing compulsory admissions for a peer-facilitated advance statement for people who had had a compulsory admission in the preceding 12 months (22).

## Aims and Objectives

The overall aim of the FINCH study was to design an intervention to reduce repeat compulsory admission, drawing on the Zürich intervention and adapting it to a UK context, and to examine the feasibility and acceptability of delivering the intervention and testing it through a randomised controlled trial.

Specific objectives were:

Phase 1: Intervention development and preliminary testing.

1. To explore through qualitative interviews (published as two separate papers (23. 24), the views of service users and clinicians on contributing factors to compulsory detention, and what kinds of interventions might prevent this.
2. To adapt and manualise the Zürich crisis planning and monitoring intervention (21) through an iterative co-design process, informed by inputs including the above qualitative interviews and relevant evidence on the implementation of crisis planning and self-management, and considering especially the needs and experiences of people from ethnic backgrounds at higher risk of detention
3. To deliver the intervention to a preliminary group of six participants, refining it further in the light of their experiences and those of clinicians delivering it.

Phase 2: Feasiblity trial

1. To assess the feasibility of delivering the intervention and investigating its effectiveness and cost-effectiveness by conducting a feasibility randomised controlled trial (RCT).
2. To assess the magnitude and direction of the difference in the risk of having at least one compulsory admission within 12 months (the proposed primary outcome for a future definitive trial) for those who were in the control arm receiving care as usual versus those in the intervention arm, and to measure potential clinical and social secondary outcomes and health economic measures.
3. To assess recruitment and retention rates for the trial measures, describe how far the intervention was received as planned, and investigate via qualitative interviews the acceptability of the intervention and of the trial measures to participating service users and clinicians.

## Methods

Full Health Research Authority (HRA) and NHS Research Ethics Committee (REC) approval was granted by the London-Bromley Research Ethics Committee (IRAS: 300671; Protocol number: 143180; REC reference: 21/LO/0734). The trial sponsor was the Joint Research Office for UCL and UCLH (Ref: 143180). The trial was pre-registered as ISRCTN11627644. Our protocol paper describes our methods in greater detail including the approach taken to developing our intervention (25).

### The Study Coproduction Group

A coproduction group was convened at the start of the study and was involved in decision making and interpretation of findings throughout. The group included people who had relevant lived experience of being detained under the Mental Health Act and/or of supporting close family and friends who had experienced this. It also included clinicians and researchers, and some members had more than one role. Our PPI lead (PN), an experienced researcher as well as an expert by experience, was a study co-applicant involved in planning the study from the initial grant application onwards. Six of our 11 Lived Experience Researchers (LERs) (including PN) were from Black ethnic backgrounds and five from White backgrounds, with a mix of ages, genders and regions of the country. Eight members of the group were clinicians, including from psychiatry, clinical psychology, occupational therapy, nursing and social work backgrounds, several also with research and/or policy backgrounds: they were also diverse in terms of ethnic group, age and gender.

### Phase 1: Intervention development

As described in more detail in our intervention development and protocol paper (25) we drew on the following sources in developing the intervention:

a. **The intervention developed and tested by Lay and colleagues in their Zürich study** (21). As well as reviewing published papers, we involved Dr Barbara Lay, lead author, as a study collaborator – she was able to describe intervention contents and experiences to us.
b. **Qualitative interviews with service users and clinician views** about what could prevent compulsory admission, conducted by the research team during the first phase of the study (23, 24).
c. **Review of Published Evidence and Interventions**: We reviewed peer-reviewed literature on effective and ineffective approaches to reducing detentions, focusing on crisis plans (12), advance statements (14), self-management for severe mental illnesses (26), and implementation of self-management interventions (27). We also reviewed a self-management and crisis planning intervention previously developed by our group in the CORE NIHR Programme (28).
d. **Guided Discussions with Co-Production Group led by relevant experts,** including on equalities and the cultural appropriateness of the intervention, and on the values and qualities that the Personal Mental Health Worker (PMHW) delivering the intervention should have.

Intervention development was guided by the Medical Research Council (MRC) Framework for Developing Complex Interventions (29, 30) and the TIDIER (Template for Intervention Description and Replication) checklist (31). The Co-Production Group met twice monthly during this phase and reviewed all inputs (summaries were presented by study researchers) and took key decisions. A manual and content for a series of sessions was developed by the study researchers and co-applicants (led by LW) and iteratively reviewed by the Co-Production Group, and by the study co-applicant team and the Personal Mental Health Workers (PMHWs) recruited to deliver the intervention, the majority of whom were clinical psychologists.

Key considerations included the centrality of therapeutic engagement, the needs for flexibility and individualisation in the intervention, including in working with different explanations of mental health and of the pathway to being compulsorily detained, and the importance of recognising and discussing impacts of culture, ethnicity and racism.

### Intervention content and delivery

The intervention consisted of four initial sessions with a PMHW, followed by monthly check-in sessions for the remainder of a year. PMHWs were clinical psychologists or other qualified mental health professionals (a mental health nurse and an occupational therapist were involved) with experience of delivering structured interventions to people with significant mental health problems. PMHWs aimed to complete as many of these initial sessions, each lasting up to an hour, as possible while participants were inpatients, and to deliver the remainder of this initial content in the community following discharge when needed.

The key tasks which PMHWs aimed to complete over four initial sessions were:

- Development of a collaborative formulation about the pathway by which the participant came to be compulsorily detained
- Creation of a personalised crisis plan, identifying early warning signs of a crisis and ways of responding, potentially including obtaining support from family, friends, and professionals. A choice of crisis plan formats was offered, including digital or paper or both. This included an opportunity to develop a “Message to future self” in written or recorded form, which could include advice on how to manage an incipient crisis or remind themselves of important aspirations.
- Development of an advanced statement recording care and treatment preferences if they became unwell in the future and lost capacity to make and communicate their wishes.
- Exploration of aspirations and recovery goals for the future.

There was considerable flexibility in the order of delivery of these activities. In the monthly check-in sessions which continued over the following year, PMHWs’ aims were to provide support, complete and revisit elements of the crisis planning intervention, and check for and prompt responses to warning signs of a crisis. The initial four sessions were mainly delivered face-to-face; phone was the main modality for check-in sessions but face to face contact, video calls and text or WhatsApp messaging were also used to maintain contact and accommodate individual preferences and circumstances.

PMHWs initially received two half-day sessions training and access to a draft manual, followed by monthly supervision with a senior clinician. They recorded details of sessions on a database, including length and location, and types of crisis management., Recording included a bespoke fidelity checklist designed to report which elements of the intervention were delivered. Participants were also invited to have therapy sessions recorded: however, given the engagement challenges in this study and feedback form our lived experience researchers, recording was presented to participants as an optional addition to the study and in practice rarely used.

### Preliminary testing

A preliminary sample of six participants meeting the eligibility criteria (see below) was recruited towards the end of the intervention development phase to receive an initial draft version of the intervention. This allowed the intervention to be studied and refined in the light of initial experiences in discussion with the Coproduction Group.

### Phase Two: Feasibility trial

#### Purpose and design

We conducted a feasibility trial, whose primary purpose was to assess the feasibility of recruitment, retention and randomisation in a trial of the intervention developed in Phase 1. We also planned to obtain the difference in risk of compulsory readmission between the experimental and control arms so as to provide the parameters for the sample size calculation of a subsequent parallel arm superiority study (compulsory readmission within a year was identified as a potential primary outcome for a full trial). An embedded qualitative study was included in order to further explore acceptability of the intervention and potential refinements ahead of a potential full trial. The trial was designed and conducted following standard guidance for feasibility trials (32,33). Online Appendix A provides a summary of the study.

#### Setting

Participants were recruited from the acute psychiatric inpatient wards of three NHS mental health Trusts in England; one in inner London, one in outer London, and one covering a mixture of non-metropolitan areas in the North-West of England. Both London Trusts have very ethnically diverse catchment areas.

#### Participants and sample size

Eligible participants were current inpatients who:

a. Had been compulsorily detained under Section 2 or Section 3 of the Mental Health Act during their current hospital admission (Section 2 allows for 28 days’ detention for assessment and Section 3 for a renewable 6-month detention).
b. Were due to receive community mental health care locally post-discharge;
c. Were aged 18 and above;
d. Had the capacity at the time of recruitment to give informed consent to participate in the trial and receive the study intervention.

Participants were excluded if they:

a. Were already receiving an intensive psychosocial intervention that focused on crisis prevention.
b. Had a diagnosis of dementia or a brain injury (no mental health conditions resulted in exclusion)
c. Did not speak sufficient English to participate without an interpreter.

We aimed to recruit 80 participants, with at least half from ethnic groups at greater risk than the general population average of being compulsorily detained, according to recent NHS data (34). These included Black African, Caribbean, British and Other, Asian Pakistani, Asian Bangladeshi and White Other^1^ ethnic groups. We aimed to recruit 30 participants from each London centre (one Inner London and one suburban Mental Health Trust) and 20 from the North-West England Trust. Our planned sample size was deemed sufficient to examine the primary aim of the study, which was to assess feasibility parameters to inform decisions about a future fully powered confirmatory trial.

The study had a statistical power of 80% (with a two-sided alpha of 5%) for detecting a reduction in the risk of compulsory readmission from 50% to 20%. Such a large difference was not considered likely, but we planned to use the true difference observed from this study to help with the future sample size calculation of a future definitive trial (if considered feasible).

#### Recruitment

Participants were recruited from acute mental health wards by methods including discussions with ward staff, screening by NHS-employed research staff, and advertising through posters and flyers on the ward. Staff considered whether patients were likely to be able to make an informed decision about study participation before introducing the study to them. Interested and potentially eligible patients were seen by a researcher and given an overview of the study and a participant information sheet, with at least 24 hours to consider participation. If they remained interested, their capacity to consent was assessed, and they were enrolled after giving written informed consent.

#### Allocation

Randomisation took place following the completion of baseline measures. Researchers at University College London who were independent of the study allocated participants via a computer-generated allocation sequence to either the intervention or control group in a 1:1 ratio using block randomisation stratified by site (3:3:2 ratio inner London: outer London: North-West England) and ethnicity (ethnic minority groups at higher risk of detention vs lower risk groups 1: 1 ratio).

Some of the study team, including the study manager (MB) remained unblinded so that they could oversee and assess intervention delivery. Other members of the study team, such as the study research assistants who were collecting follow-up outcomes, were kept blind to treatment allocation as far as possible.

#### Interventions

Participants randomised to the intervention group received the co-designed intervention, delivered as above by a PMHW, alongside usual acute and post-discharge care. Control group participants received usual care only, usually involving discharge to the care of a generic community mental health team, or a specialist team such as those supporting people with early intervention in psychosis or a personality disorder diagnosis.

#### Measures

##### Descriptive data

Demographic data from participants and casenote data on diagnosis were collected at baseline to characterise the sample.

##### Outcomes

The primary goal was to assess feasibility outcomes. Data on candidate outcomes for a future randomised controlled trial of the intervention were also collected, allowing assessment of the feasibility and best methods of collecting these outcomes, collection of data to inform a power calculation for a definitive trial, and a preliminary assessment of the likelihood of a positive result from a definitive trial.

##### Feasibility Outcomes

Feasibility parameters included recruitment rates, acceptance of randomisation, rates and patterns of attrition from treatment and trial assessments, delivery of each intervention component, completion rates for individual outcome measures, and rates of serious adverse events in each arm of the trial.

##### Candidate trial outcomes

Candidate trial outcomes were measured via clinical records and interviews. Research interviews were conducted by the research team at baseline prior to randomisation, and at 6 and 12 months after randomisation. A final follow-up point at 24 months (which will be reported elsewhere) involves health record data only.

##### Primary Trial Outcome

The planned primary outcome for a future definitive trial was whether the participant had been compulsorily detained in hospital under Section 2 or Section 3 of the Mental Health Act within one year of randomisation.

##### Secondary outcomes

The following candidate secondary outcomes were assessed at interview by a researcher at baseline and at 6 and 12 month follow up where consent was given to a further interview:

a. Satisfaction with services was examined using the Client Satisfaction Questionnaire (CSQ) (35). This is an 8-item scale where participants can rate their satisfaction with various aspects of their care on a 4-point Likert scale.
b. Self-rated recovery was measured by the 15-item Questionnaire about the Process of Recovery (QPR) (36). Participants can score from 0 (disagree strongly) to 4 (agree strongly) on each item and score up to a maximum of 60 on the scale.
c. Self-management confidence was measured using the Mental Health Confidence scale (37). Participants report their confidence in managing their mental health for 16 items rated on a Likert scale from very non-confident to very confident.
d. Health-related quality of life was measured by the REQOL-10 (38) and EQ-5D-5L (39). Both measures are used to derive quality-adjusted life years (QALYs). The REQOL is specifically designed for mental health studies and participants rate their quality of life on 10 items from 0 to 4. The EQ-5D-5L, is a generic measure and more commonly used. It contains five domains (mobility, self care, usual activities, pain/discomfort, and anxiety/depression). With each rated between 1 (no problems) to 5 (extreme problems).. Both the REQOL and EQ-5D-5L are converted to a weight with 1 representing full health. This weight is subsequently used to derive QALYs.
e. Psychiatric symptoms were assessed using the Brief Psychiatric Rating Scale (BPRS) (40). Participants were rated by a researcher on 18 items from 0/ NA (not assessed) to 7 (extremely severe) to give an overall score for psychiatric symptoms.
f. Service use data were collected using an adapted version of the Client Service Receipt Inventory (CSRI) (41). Costs were calculated by combining this information with appropriate unit costs. The costs of the intervention were calculated from information on staff time and other requirements. Intervention costs were based on the number of therapy sessions delivered and the unit cost of these.

##### Analysis

The following steps were carried out, adhering to an analysis plan published online ahead of analyses being conducted (42).

##### Descriptive Statistics

Baseline characteristics were summarised for all patients in the study. Summary measures for the baseline characteristics were presented as mean and standard deviation for continuous (approximate) normally distributed variables, medians and interquartile ranges for non-normally distributed continuous variables, and frequencies and percentages for categorical variables.

##### Feasibiility outcomes: recruitment, retention and intervention delivery

Recruitment rate, retention in experimental and control groups and rates of collection of candidate primary and secondary outcome measures and intervention delivery parameters were also summarised descriptively. Progression criteria were established for consideration of whether progress to a full trial was warranted (26). These were that achieving the goals below in full would indicate trial protocols and procedures were suitable for a full trial, while falling short by up to 20% on any parameter would indicate a need to consider what improvements could be made to make a full trial achievable, and falling short by more than 20% would suggest a need for major changes and a further feasibility trial if a full trial were to be conducted:

- Recruitment within 9 months of 80 trial participants.
- At least 50% of these participants are from ethnic backgrounds associated with an elevated risk of being compulsorily admitted.
- At least 85% data completeness on primary outcome measure for trial (repeat compulsory admission within a year)
- At least 60% data completeness for secondary outcomes at 1 year
- At least 75% of intervention participants to have developed a crisis plan and have received at least 3 intervention sessions (our study definition of receiving minimum *per protocol* treatment).

##### Analysis of candidate primary outcome for a trial

This feasibility study did not have sufficient statistical power to assess the effectiveness of the intervention with precision but allowed an assessment of whether the direction and magnitude of any effect found for the proposed primary outcome are consistent with a hypothesis that the programme is effective in reducing repeat detentions. For this reason and to test the analysis envisaged for a future, fully powered, effectiveness RCT, the primary outcome at follow-up was compared between study arms using appropriate multi-level models. Hierarchical multi-level modelling, allowing for clustering of residuals between centres by introducing the variable coding for centres as a random intercept was utilised. We entered all other stratification variables as fixed effects. This analysis was also conducted for those in ethnic groups at higher risk of compulsory admission alone.

##### Adverse Events

Adverse events were summarised in terms of the number of (serious) adverse events and the number of participants with any (serious) adverse events in each randomised group and compared using Fisher’s exact test.

### Phase Two Qualitative Study to investigate experiences of receiving and delivering the intervention

#### Sample

We aimed to interview up to 20 participants who were allocated to receive the FINCH intervention, as well as the PMHWs at each site who delivered the intervention. If a choice of participants were available, we aimed to use purposive sampling to ensure a full range of demographic characteristics and service experiences were included.

#### Interview guide

The interview topic guide for participants allocated to receive the FINCH intervention was developed collaboratively by the Co-Production Group and the FINCH research team. The core research team developed the interview topic guide for the PMHWs delivering the intervention. For each group, we aimed to explore experiences of intervention delivery, most and least helpful elements, and perceived influence on recovery. Online Appendix B shows the topic guides.

Interviews were conducted between November 2022 and January 2024. Capacity was assessed and informed consent recorded both to conduct and record the interview. Informed consent was sought for all interviews to audio or video record them and have them transcribed verbatim by an external transcription agency approved as secure by UCL. All audio-transcripts were then checked by the researchers and any identifying information was anonymised. Interviews with participants allocated to receive the FINCH intervention were conducted by an LER from a Co-Production Group who had received training in qualitative interviewing and those with clinicians by FINCH researchers.

Interviews were analysed deductively using Framework Analysis (24), a form of thematic analysis. The framework consisted of themes derived from the seven constructs of the Theoretical Framework of Acceptability of Healthcare Interventions(39), which are affective attitude, burden, intervention coherence, ethicality, opportunity costs, perceived effectiveness, and self-efficacy.

## Results

### Trial findings

Feasibility outcomes

### Recruitment

Eighty participants were recruited and gave informed consent to participate in the trial: 30 from each participating London Trust and 20 from the northern Trust. Recruitment took place over a 9-month period, as had been specified in the trial protocol and the planned sample was achieved for each participating Trust.

### Randomisation

Thirty-eight participants were randomly assigned to receive the intervention and 42 to the control condition. Figure 1 (the Consort diagram) shows flow through the study. It should be noted that the 590 identified as potential participants were a minority of potentially eligible inpatients compulsorily detained in the participating wards during the study period: recruitment on these wards took place intermittently, reflecting the availability of researchers and of PMHWs who could deliver the intervention.

**Figure 1:**
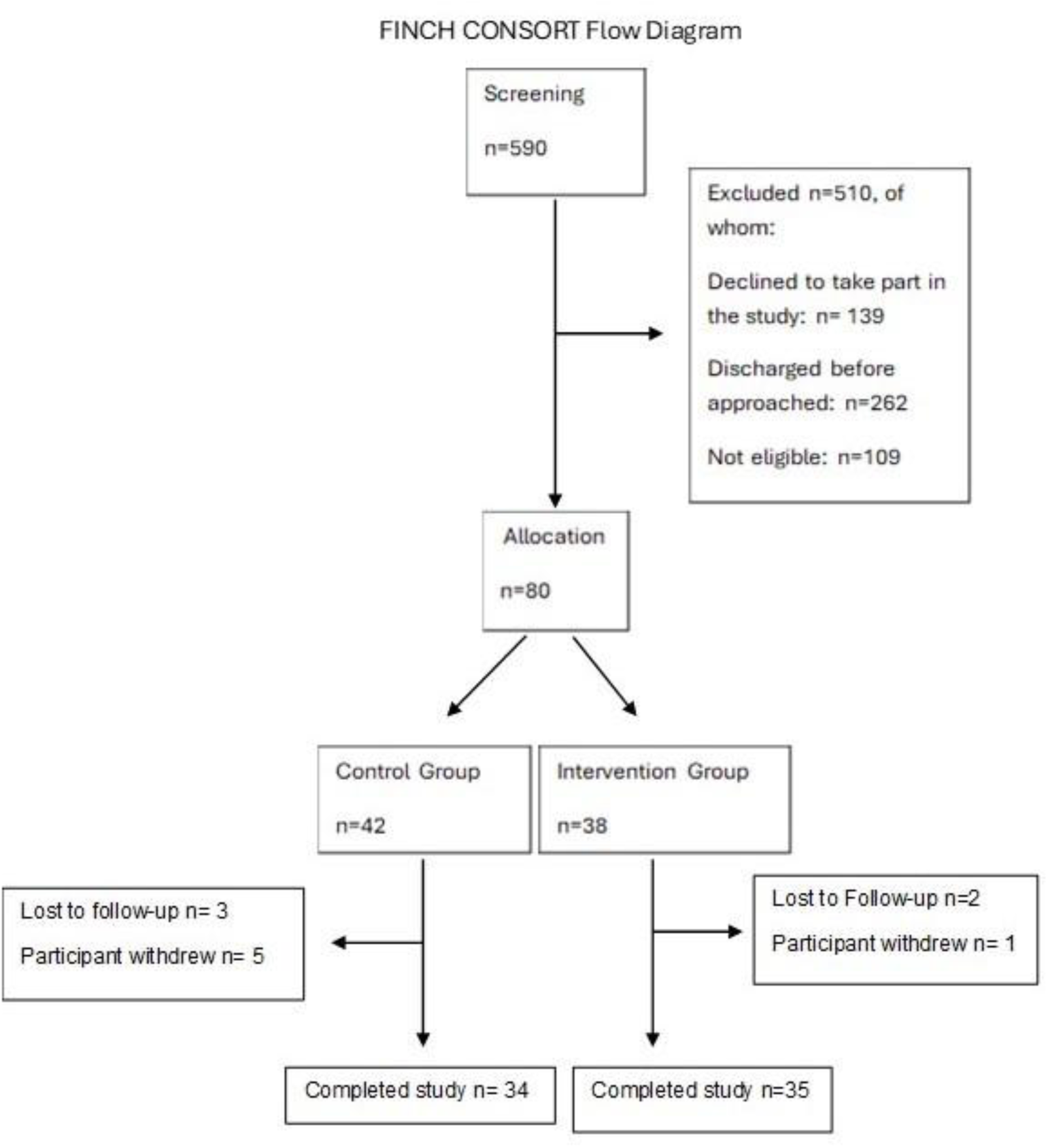
Flow diagram

### Sample characteristics

Table 1 shows sample characteristics. Two participants asked for all data to be withdrawn and are shown as missing in the table below.

**Table 1:**
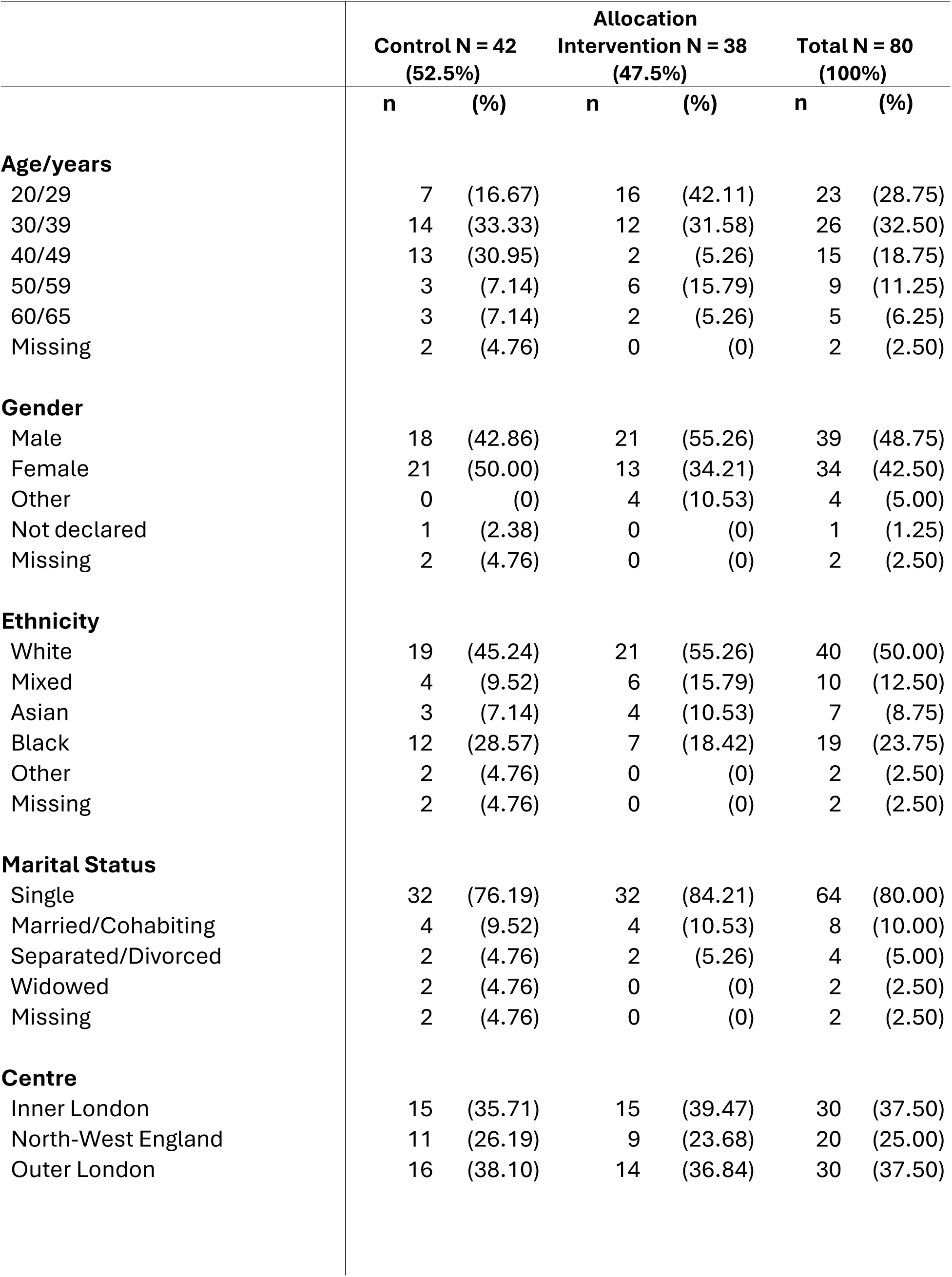
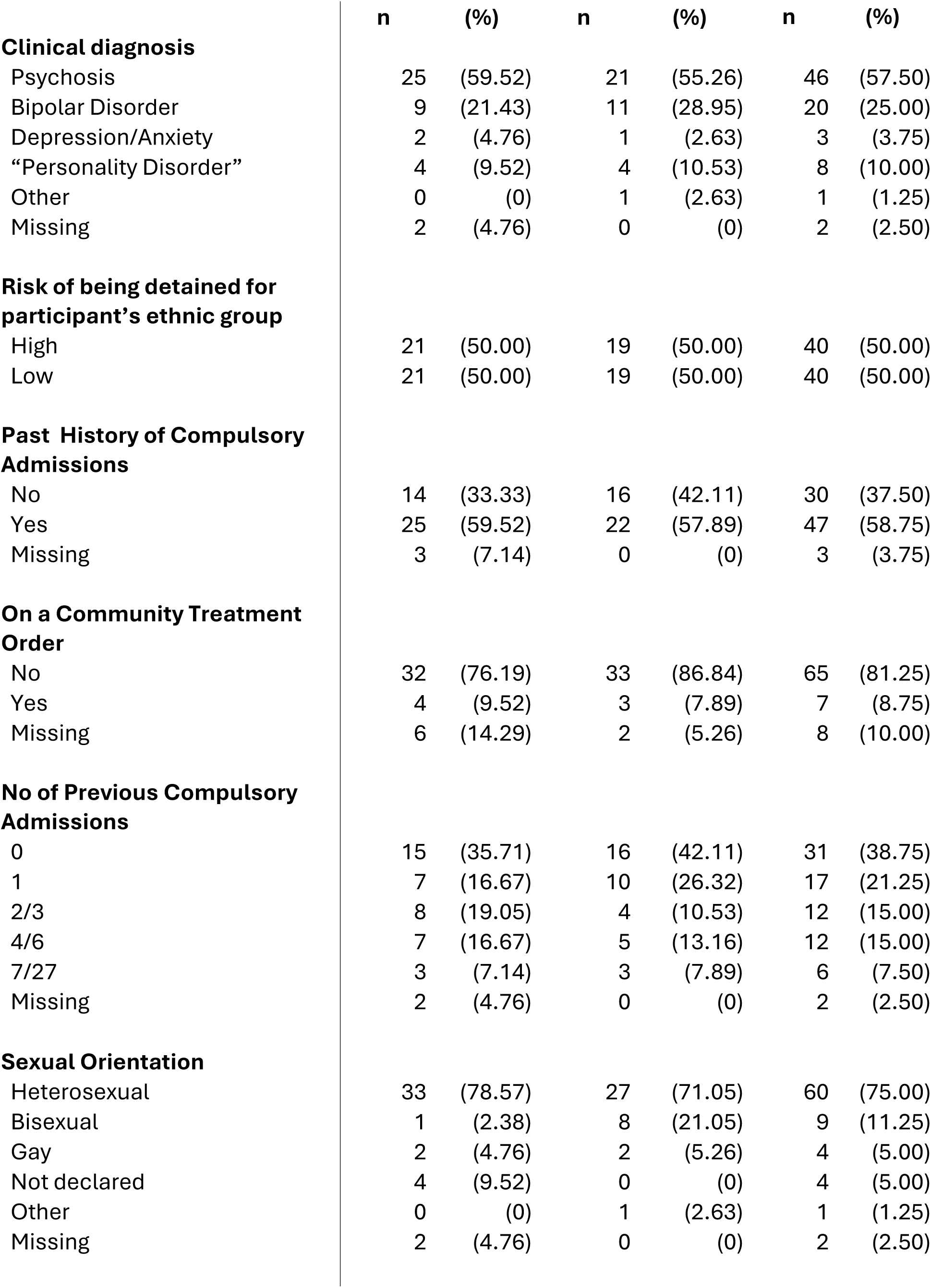
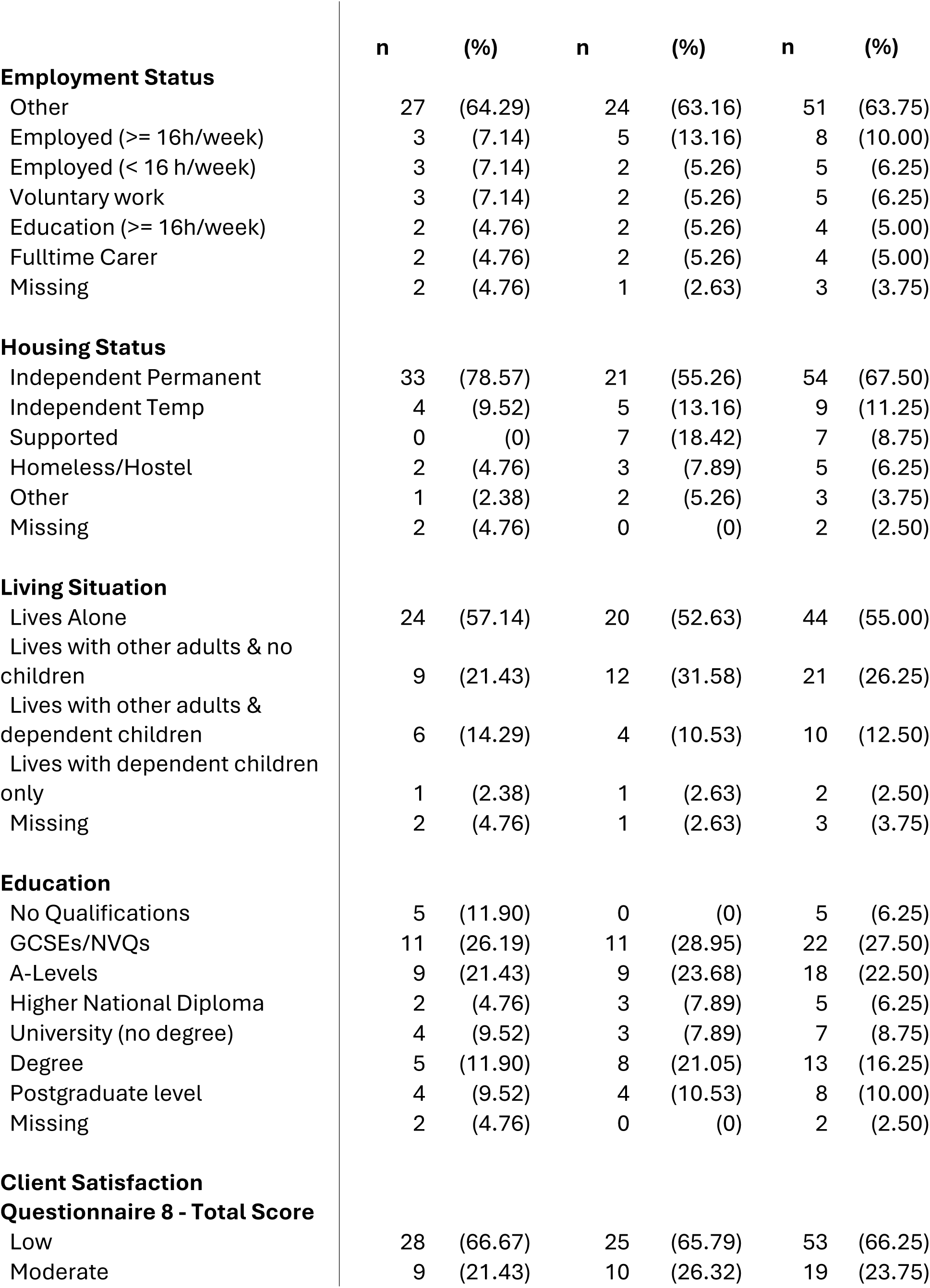
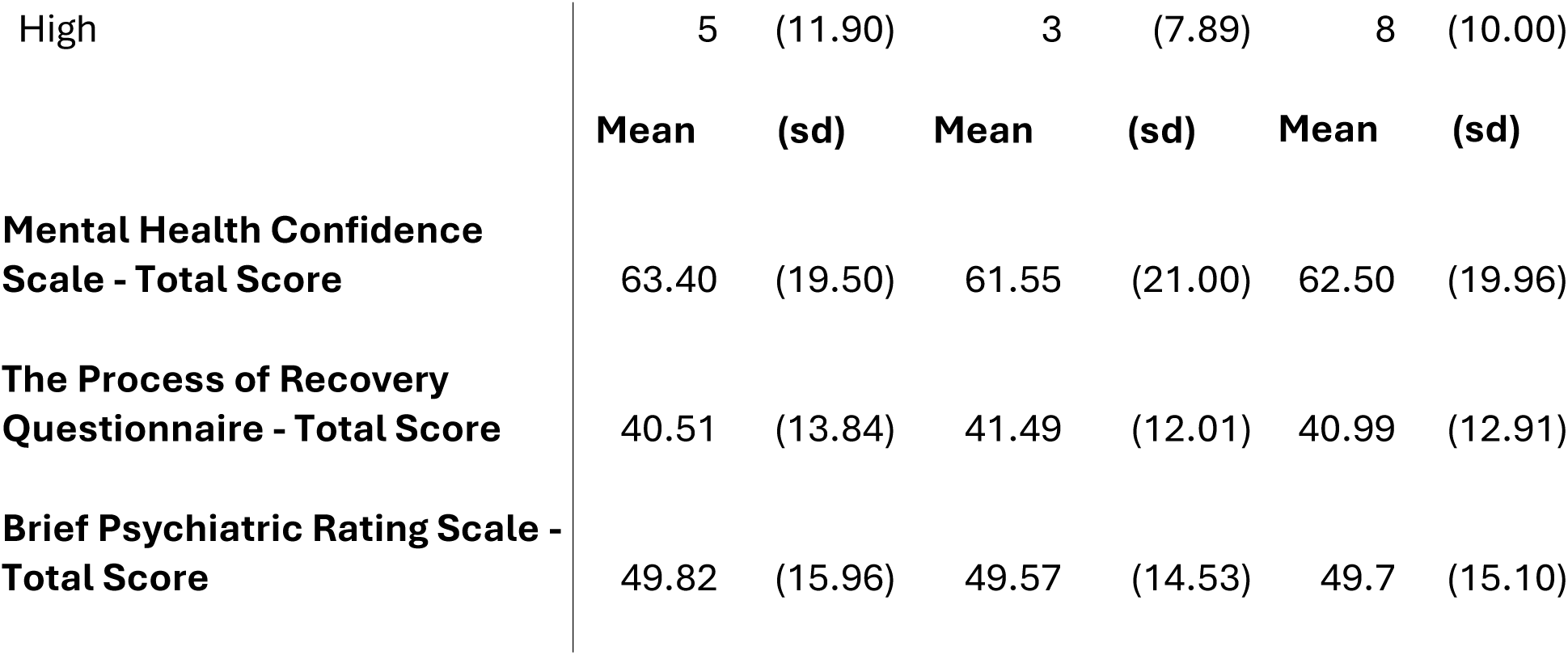
Baseline characteristics of intervention and control groups.

|  | Allocation |  |  |  |  |  |
| --- | --- | --- | --- | --- | --- | --- |
|  | Control N = 42<br>(52.5%) |  | Intervention N = 38<br>(47.5%) |  | Total N = 80<br>(100%) |  |
|  | n | (%) | n | (%) | n | (%) |
| **Age/years** |  |  |  |  |  |  |
| 20/29 | 7 | (16.67) | 16 | (42.11) | 23 | (28.75) |
| 30/39 | 14 | (33.33) | 12 | (31.58) | 26 | (32.50) |
| 40/49 | 13 | (30.95) | 2 | (5.26) | 15 | (18.75) |
| 50/59 | 3 | (7.14) | 6 | (15.79) | 9 | (11.25) |
| 60/65 | 3 | (7.14) | 2 | (5.26) | 5 | (6.25) |
| Missing | 2 | (4.76) | 0 | (0) | 2 | (2.50) |
| **Gender** |  |  |  |  |  |  |
| Male | 18 | (42.86) | 21 | (55.26) | 39 | (48.75) |
| Female | 21 | (50.00) | 13 | (34.21) | 34 | (42.50) |
| Other | 0 | (0) | 4 | (10.53) | 4 | (5.00) |
| Not declared | 1 | (2.38) | 0 | (0) | 1 | (1.25) |
| Missing | 2 | (4.76) | 0 | (0) | 2 | (2.50) |
| **Ethnicity** |  |  |  |  |  |  |
| White | 19 | (45.24) | 21 | (55.26) | 40 | (50.00) |
| Mixed | 4 | (9.52) | 6 | (15.79) | 10 | (12.50) |
| Asian | 3 | (7.14) | 4 | (10.53) | 7 | (8.75) |
| Black | 12 | (28.57) | 7 | (18.42) | 19 | (23.75) |
| Other | 2 | (4.76) | 0 | (0) | 2 | (2.50) |
| Missing | 2 | (4.76) | 0 | (0) | 2 | (2.50) |
| **Marital Status** |  |  |  |  |  |  |
| Single | 32 | (76.19) | 32 | (84.21) | 64 | (80.00) |
| Married/Cohabiting | 4 | (9.52) | 4 | (10.53) | 8 | (10.00) |
| Separated/Divorced | 2 | (4.76) | 2 | (5.26) | 4 | (5.00) |
| Widowed | 2 | (4.76) | 0 | (0) | 2 | (2.50) |
| Missing | 2 | (4.76) | 0 | (0) | 2 | (2.50) |
| **Centre** |  |  |  |  |  |  |
| Inner London | 15 | (35.71) | 15 | (39.47) | 30 | (37.50) |
| North-West England | 11 | (26.19) | 9 | (23.68) | 20 | (25.00) |
| Outer London | 16 | (38.10) | 14 | (36.84) | 30 | (37.50) |

|  | n | (%) | n | (%) | n | (%) |
| --- | --- | --- | --- | --- | --- | --- |
| **Clinical diagnosis** |  |  |  |  |  |  |
| Psychosis | 25 | (59.52) | 21 | (55.26) | 46 | (57.50) |
| Bipolar Disorder | 9 | (21.43) | 11 | (28.95) | 20 | (25.00) |
| Depression/Anxiety | 2 | (4.76) | 1 | (2.63) | 3 | (3.75) |
| “Personality Disorder” | 4 | (9.52) | 4 | (10.53) | 8 | (10.00) |
| Other | 0 | (0) | 1 | (2.63) | 1 | (1.25) |
| Missing | 2 | (4.76) | 0 | (0) | 2 | (2.50) |
| **Risk of being detained for participant’s ethnic group** |  |  |  |  |  |  |
| High | 21 | (50.00) | 19 | (50.00) | 40 | (50.00) |
| Low | 21 | (50.00) | 19 | (50.00) | 40 | (50.00) |
| **Past History of Compulsory Admissions** |  |  |  |  |  |  |
| No | 14 | (33.33) | 16 | (42.11) | 30 | (37.50) |
| Yes | 25 | (59.52) | 22 | (57.89) | 47 | (58.75) |
| Missing | 3 | (7.14) | 0 | (0) | 3 | (3.75) |
| **On a Community Treatment Order** |  |  |  |  |  |  |
| No | 32 | (76.19) | 33 | (86.84) | 65 | (81.25) |
| Yes | 4 | (9.52) | 3 | (7.89) | 7 | (8.75) |
| Missing | 6 | (14.29) | 2 | (5.26) | 8 | (10.00) |
| **No of Previous Compulsory Admissions** |  |  |  |  |  |  |
| 0 | 15 | (35.71) | 16 | (42.11) | 31 | (38.75) |
| 1 | 7 | (16.67) | 10 | (26.32) | 17 | (21.25) |
| 2/3 | 8 | (19.05) | 4 | (10.53) | 12 | (15.00) |
| 4/6 | 7 | (16.67) | 5 | (13.16) | 12 | (15.00) |
| 7/27 | 3 | (7.14) | 3 | (7.89) | 6 | (7.50) |
| Missing | 2 | (4.76) | 0 | (0) | 2 | (2.50) |
| **Sexual Orientation** |  |  |  |  |  |  |
| Heterosexual | 33 | (78.57) | 27 | (71.05) | 60 | (75.00) |
| Bisexual | 1 | (2.38) | 8 | (21.05) | 9 | (11.25) |
| Gay | 2 | (4.76) | 2 | (5.26) | 4 | (5.00) |
| Not declared | 4 | (9.52) | 0 | (0) | 4 | (5.00) |
| Other | 0 | (0) | 1 | (2.63) | 1 | (1.25) |
| Missing | 2 | (4.76) | 0 | (0) | 2 | (2.50) |

**Employment Status**
|  | n | (%) | n | (%) | n | (%) |
| --- | --- | --- | --- | --- | --- | --- |
| Other | 27 | (64.29) | 24 | (63.16) | 51 | (63.75) |
| Employed (>= 16h/week) | 3 | (7.14) | 5 | (13.16) | 8 | (10.00) |
| Employed (< 16 h/week) | 3 | (7.14) | 2 | (5.26) | 5 | (6.25) |
| Voluntary work | 3 | (7.14) | 2 | (5.26) | 5 | (6.25) |
| Education (>= 16h/week) | 2 | (4.76) | 2 | (5.26) | 4 | (5.00) |
| Fulltime Carer | 2 | (4.76) | 2 | (5.26) | 4 | (5.00) |
| Missing | 2 | (4.76) | 1 | (2.63) | 3 | (3.75) |
| **Housing Status** |  |  |  |  |  |  |
| Independent Permanent | 33 | (78.57) | 21 | (55.26) | 54 | (67.50) |
| Independent Temp | 4 | (9.52) | 5 | (13.16) | 9 | (11.25) |
| Supported | 0 | (0) | 7 | (18.42) | 7 | (8.75) |
| Homeless/Hostel | 2 | (4.76) | 3 | (7.89) | 5 | (6.25) |
| Other | 1 | (2.38) | 2 | (5.26) | 3 | (3.75) |
| Missing | 2 | (4.76) | 0 | (0) | 2 | (2.50) |
| **Living Situation** |  |  |  |  |  |  |
| Lives Alone | 24 | (57.14) | 20 | (52.63) | 44 | (55.00) |
| Lives with other adults & no children | 9 | (21.43) | 12 | (31.58) | 21 | (26.25) |
| Lives with other adults & dependent children | 6 | (14.29) | 4 | (10.53) | 10 | (12.50) |
| Lives with dependent children only | 1 | (2.38) | 1 | (2.63) | 2 | (2.50) |
| Missing | 2 | (4.76) | 1 | (2.63) | 3 | (3.75) |
| **Education** |  |  |  |  |  |  |
| No Qualifications | 5 | (11.90) | 0 | (0) | 5 | (6.25) |
| GCSEs/NVQs | 11 | (26.19) | 11 | (28.95) | 22 | (27.50) |
| A-Levels | 9 | (21.43) | 9 | (23.68) | 18 | (22.50) |
| Higher National Diploma | 2 | (4.76) | 3 | (7.89) | 5 | (6.25) |
| University (no degree) | 4 | (9.52) | 3 | (7.89) | 7 | (8.75) |
| Degree | 5 | (11.90) | 8 | (21.05) | 13 | (16.25) |
| Postgraduate level | 4 | (9.52) | 4 | (10.53) | 8 | (10.00) |
| Missing | 2 | (4.76) | 0 | (0) | 2 | (2.50) |
| **Client Satisfaction****Questionnaire 8 - Total Score** |  |  |  |  |  |  |
| Low | 28 | (66.67) | 25 | (65.79) | 53 | (66.25) |
| Moderate | 9 | (21.43) | 10 | (26.32) | 19 | (23.75) |
| High | 5 | (11.90) | 3 | (7.89) | 8 | (10.00) |
|  | **Mean** | **(sd)** | **Mean** | **(sd)** | **Mean** | **(sd)** |
| **Mental Health Confidence Scale - Total Score** | 63.40 | (19.50) | 61.55 | (21.00) | 62.50 | (19.96) |
| **The Process of Recovery Questionnaire - Total Score** | 40.51 | (13.84) | 41.49 | (12.01) | 40.99 | (12.91) |
| **Brief Psychiatric Rating Scale - Total Score** | 49.82 | (15.96) | 49.57 | (14.53) | 49.7 | (15.10) |

A feasibility goal for the study was to recruit at least half participants from groups at elevated risk of compulsory admission, including Black or Black British, Asian Bangladeshi or Pakistani, White Other and some Mixed groups. Forty of the 80 participants fell into this category.

### Completion of candidate measures for a full RCT

#### Proposed primary outcome for trial

Data were collected for the candidate primary outcome for a full trial, whether participants had been compulsorily detained again by the one-year follow-up point for 69/80 (86%) of participants. Six participants withdrew from the study and asked that further data not be collected from their clinical records (two also asked us to remove baseline data). A further five participants moved out of the catchment areas in which the study was conducted and we were unable to obtain data on subsequent service use.

#### Secondary measures

At six-months follow-up, secondary outcome data were collected on interview measures for 46/80 participants (58%). Among the 34 with no follow-up data, 17 participants declined to provide follow-up data, 13 were not contactable after several attempts, 3 were unwell/readmitted at time of follow-up, and one participant was felt not to be suitable for follow-up as recovering from a distressing experience (not study-related).

At 12 months, secondary outcome data were collected for 41/80 participants (51%). Of the 39 not interviewed, 21 declined to provide follow-up data for researchers, 14 were not contactable after several attempts, of whom 3 had moved out of area, and 4 appeared to be too unwell to participate in an interview.

#### Intervention Delivery

Initially, seven clinicians, two in two centres, and three in the third centre, were employed as PMHWs to deliver the FINCH intervention and were provided the draft manual and a one day training in delivering the intervention. The PMHWs were six clinical psychologists, three of whom worked on adult acute inpatient wards, and an occupational therapist. Four left the role before the planned end date (including two for maternity leave), following which a further three clinicians, a psychiatric nurse and two clinical psychologists, were recruited to continue delivering the intervention. Several participants experienced a delay in starting intervention the intervention due to lack of PMWH availability or a change of PMHW towards the end of intervention delivery. It was noted that the relatively small time allocations (often just a half day a week) for working on this feasibility trial seemed to make it difficult to recruit clinicians to this role and for them to be sufficiently flexible in how they delivered the intervention. Our manual recommended that the initial four sessions be delivered as far as possible while participants were still inpatients, with several in a week if feasible and acceptable. However, in practice PMHWs’ limited available time meant that most participants received sessions only weekly, with the initial four sessions often continuing after discharge.

#### Receipt of intervention

Of the 38 participants allocated to receive the FINCH intervention, six did not start. Three of these participants moved out of area on discharge following randomisation (despite the research team seeking to exclude people who were about to move) and three were quickly discharged from the ward following randomisation and did not subsequently respond to multiple attempts to contact them.

Of the remaining 32 participants allocated to the intervention group, seven participants asked to withdraw from receiving the intervention at varying stages during its delivery: three of these had not yet received the four initial sessions, while the other four had received all initial sessions and up to eight follow-up contacts. Thirty-two participants attended the first session, after which two participants were lost to follow-up following hospital discharge having received only one session. Twenty five of the 38 intervention group participants (66%) received at least three intervention sessions and had at least partially developed a written crisis plan: this was the pre-specified minimum for participants to be classified as having received the intervention. Fifteen of these participants were from ethnic groups at high risk of being detained under the MHA.

#### Delivery of intervention components

Table 2 shows which components of the planned intervention were delivered. PMHWs were encouraged to prioritise completion of crisis plans within available time.

**Table 2.** Delivery of components of the intervention.

| . | Formulation completed about how crisis developed | Crisis plan developed | Advanced statement recorded | Recovery goals recorded |
| --- | --- | --- | --- | --- |
| Completed | 24 | 22 | 14 | 11 |
| Partially completed | 5 | 5 | 1 | 9 |
| Declined to complete | 0 | 1 | 5 | 1 |
| *Did not complete but not declined | 3 | 4 | 12 | 11 |

\* excludes six participants who did not start intervention

##### Follow-up sessions

For those who participated in at least one follow-up check in session, the median number of follow-up contacts was four, with the total number ranging from one to ten. Sessions did not reliably take place on a monthly basis as in the protocol for reasons related to both participant and clinician availability.

##### Serious adverse events

Table 3 shows Serious Adverse Events (SAEs) in each arm, identified from PMHW reports and from case notes. Almost all SAEs were hospital admissions and were not suspected to be study-related. One SAE was discussed with the sponsor and independent chair of the Trial Steering Committee as possibly study-related, but the conclusion from these discussions was that this was unlikely.

**Table 3:** Serious Adverse Events by arm of study.

|  | Data available for full period | Number of participants with SAEs | Total number of SAEs |
| --- | --- | --- | --- |
| Intervention group | n=35/38 | 11 | 17<br>-Psychiatric hospital admission: 11<br>-General hospital admission: 6 |
| Control Group | n=34/42 | 14 | 25<br>-Psychiatric hospital admission: 17<br>-General Hospital admission: 8 |

#### Findings on candidate trial primary outcome measure

Regarding the candidate primary outcome measure for a full trial, at 12 months’ follow-up from recruitment, 49 of the 69 (71.0%) for whom data were available on the primary outcome had not been compulsorily detained again. Twenty-three (67.6%) of those allocated to the control group for whom follow-up data were available had not been detained again during the year after baseline, while 26 (74.3%) of those who had been allocated to the intervention group had not been detained again. Those who received the intervention had an increased odds ratio of avoiding detention of 1.378 (95% Confidence Interval: 0.480 to 3.955) versus those randomly allocated to the control arm: the number needed to treat to prevent one detention was estimated as sixteen.

When only members of ethnic groups at a high risk of compulsory admission were included, at 12 months follow-up, 10 (58.8%) of those who had been allocated to the control group and had data available had not been detained vs. 12 (70.6%) of those who been allocated to the intervention group (Table 5). This corresponded to increased odds of not being detained of 1.680; (95% CI: 0.405 to 6.962) for the group receiving the intervention. Number needed to treat within this sub-group to prevent one detention was estimated as nine. No findings reached the threshold for statistical significance, as anticipated with the small sample size and very limited power in this feasibility trial.

No analysis involving diagnosis was planned: however, given different rates in the general population, members of the research team raised the question *post hoc* of whether patterns of diagnosis were markedly different between high risk and low risk ethnic groups. In the high risk ethnic groups, 36/39 of those with an available casenote diagnosis had psychosis or bipolar, compared with 30/39 in the low risk group.

Further details regarding distribution of the primary outcome, including risk differences between arms are found in **Online Appendix C (Tables C1 and C2).**

#### Proposed trial secondary measures

**Online Appendix D** shows findings for the candidate secondary measures for a full trial that were collected at interview. No statistical analyses beyond descriptive statistics were planned for these measures given the status of the trial as a feasibility study. It should also be noted that there was considerable attrition for these measures.

#### Health economic findings

**Online Appendix E** presents the health economic findings in full. Costs were measured using the Client Service Receipt Inventory (CSRI) (38) and covered the six-month periods prior to baseline and 6- and 12-month follow-up, encompassing primary and secondary care and social care services. Mean total costs for the combined follow-up period are compared with bootstrapped 95% confidence intervals generated around the cost difference between the two groups. We included the EQ-5D-5L (36) as a measure of health-related quality of life to be used in health economic analyses, although as with all interview secondary measures there was substantial attrition on this measure.

Total costs could be computed from the CSRI for 40 (95.2%) control group and 36 (94.7%) intervention group participants at baseline, 36 (85.7%) control group and 37 (97.4%) intervention group participants at 6-month follow-up, and 29 (69.0%) control group and 34 (89.5%) intervention group participants at 12-month follow-up.

All participants had inpatient stays in the six months prior to baseline, and most also in the first six months of follow up, as discharge did not usually occur immediately after randomisation. In the six months prior to the 12-month follow-up point there were fewer in the intervention group who had inpatient stays. Use of GPs was slightly more likely for the intervention group prior to baseline, but prior to the 12-month follow-up this had switched with more of the usual care group seeing a GP. Contacts with psychiatrists, mental health nurses, and other mental health workers were common for both groups in each period but without major differences between groups.

The mean (SD) cost for the FINCH study intervention was £583 (£404) for the intervention group, with a range of £0 to £1385. Other care costs were dominated by inpatient costs. The total mean (SD) cost over the combined follow-up period, including the intervention, was £41,840 (£29,068) for the control group and £35,962 (£38,020) for the intervention group. The mean incremental cost was −£5872 (i.e. lower for the intervention group) with a bootstrapped 95% confidence interval of −£22,204 to £9781, showing the difference not to be statistically significant.

### Phase Two Qualitative Study Findings

#### Qualitative interviews to investigate experiences of receiving and delivering the intervention

Informed consent to participate in a recorded interview regarding experiences of receiving and delivering the intervention was obtained within the timeframe available from eight service users who had received the intervention, and nine of the PMHWs who had delivered it. If a larger number of service user participants had been available, we would have sampled purposively to ensure inclusion of the main demographic and diagnostic groups participating in the study, but limited numbers meant we included all who consented. These data were analysed in relation to the categories in the Theoretical Framework of Acceptability, which provides a structured approach to exploring the main aspects of acceptability (43). The following is a brief summary: a more detailed description of the sample and analysis will be published separately.

#### Affective Attitude

This construct assesses overall perceptions of the intervention and its content. Positive comments from service users included feeling supported and able to express their views openly, gaining a greater understanding of their illness, and benefiting from the crisis planning process.

> “It has been a positive experience because it has put my… put where I’m at in my illness to some sort of reality so to speak. I don’t know whether that makes sense. It has made me get more insight into my illness basically.” (Service User Participant)

The opportunity to form a strong and consistent relationship with PMHWs was also valued, with some seeing PMHWs as akin to “friends” or “peers,” with less than usual power imbalance. PMHWs also reported a positive overall view of the intervention, finding the manual content useful and the opportunity to spend time forming a therapeutic relationship at this important stage valuable. Suggestions for improvement included offering the intervention sooner so that a larger proportion was delivered on the inpatient ward, and more frequent follow-up sessions (although few in practice took up all that were offered).

### Burden

This construct examines the effort required for intervention engagement. Most service users found the intervention manageable, although one reported a substantial interruption due to staff unavailability and another financial barriers to participating. PMHWs saw the intervention as time-consuming, especially because of the efforts needed to keep in touch and engage with service users following discharge. The time allocated for the intervention in the study was seen by most as insufficient for the persistent efforts required:

> “It’s been frustrating… I think it’s been quite time consuming. Making some space to see someone and then they don’t turn up, then chasing to see if they’re okay. If a phone call doesn’t work, sending a text and putting lots of reminders in my calendar, ‘Contact this person if they don’t reply in a week.’ There are a couple I tried. One said they weren’t interested and one just never really…” - (PMHW Participant)

### Ethicality

This construct focuses on how well the intervention aligns with the participants’ values. The moral and ethical consequences of the intervention were not discussed by many participants. One participant reported that the intervention made them feel as though they mattered, whilst another felt that the intervention would only be useful for those wishing to recover.

> “It was something that I wasn’t expecting that is good. To be completely honest, I don’t believe that it would work on everybody. I think that it has to be individual on a certain level of capacity to appreciate those sessions, somebody who wants to recover. It cannot be somebody who is ready to fall into problems again.” (Service User Participant)

### Intervention coherence

This describes the extent to which participants understand the intervention and how it is intended to work. Both PMHWs and service users felt they had a good understanding of the purpose and content of the intervention:

> “Building a crisis plan and increasing self-awareness…” (Service User Participant)

> “it gave me the tools that I needed at the time and now to… know when I’m having episodes or… put things in place where it won’t go to that level of going back to hospital, hopefully.” (Service User Participant)

> “ it does focus on their experience of being sectioned, the reasons why they’ve come into hospital and what can be done to support them before them being re-admitted.” (PMHW Participant)

### Opportunity costs

This describes the extent to which benefits, profits, or values must be given up to engage in the intervention. Interviewed service users did not describe such opportunity costs. PMHWs, for whom in most cases delivering the intervention was a relatively small part of their overall job role, found it difficult to fit in, especially if they were persistent and flexible in making contact with service users, as the intervention manual requires:

> “Obviously, trying to work this around I’ve got a full diary of things that I need to do. It was scheduled at a time where… I’m doing this on the bank, I’m not doing this as part of my job. That has been a bit challenging. If I’m attending that in work time, I have to work it back.” (PMHW Participant)

Attending the monthly group clinical supervision sessions also was challenging due to other clinical roles.

### Perceived effectiveness

This describes how far the intervention is seen as likely to achieve its purpose. Interviewed service users described changes in their self-management strategies following receiving the intervention:

> “I’ve noticed I’m more aware of my triggers. And the choices I’m making now. Like don’t drink, don’t take any illicit drugs, like weed and stuff, because I used to think that was helping me, but really and truly it wasn’t helping.” (Service User Participant)

Clinicians delivering the intervention also believed that the intervention had potential to achieve its purpose:

> “Actually, going through some of things that others can do, what they can do, thinking about the signs and triggers. It feels to me that it’s been quite helpful for them. The few who have engaged really well have said it’s not something that they’ve done before and it’s quite helpful to have.” (PMHW Participant)

### Self efficacy

This construct captures how far participants are confident that they can carry out their role in the intervention. Clinicians were confident in their ability to deliver the intervention after the training, provided they could engage participants.

> “it was really helpful to get an overview of the intervention. And then, the supervision sessions have been really helpful in terms of if any questions have come up, or thinking about having to adapt it and tailor it a bit more, then that’s been really supportive, to think more about it there as well.” (PMHW Participant)

Some of those who received the intervention reported having acquired skills that would enable them to mobilize self-management skills better in future:

> “I think I could make my own crisis plan now if I needed to. I understand, which is something I used to find overwhelming. In fact, I found doing crisis plans very difficult because then I wouldn’t be able to think of things. So I feel like I’ve learnt how to do crisis plan better.” (Service User Participant),

## Discussion

### Main findings

The main aim of this study was to assess the feasibility and acceptability of delivering and evaluating through a randomised controlled trial a co-designed crisis planning intervention. We published a set of progression criteria in our protocol paper (25), indicating that progress to a full trial would be fully justified if these criteria were met in full, and that a shortfall of less than 20% would indicate potential to progress to a full trial after reviewing and addressing difficulties identified in the feasibility trial. Our feasibility trial’s performance in relation to these progression criteria was as follows:

**Recruitment within 9 months for 80 trial participants:** this was achieved within the acute wards of the intended three catchment areas.

**At least 50% of these participants are from ethnic backgrounds associated with an elevated risk of being compulsorily admitted:** this was also achieved, with half the participants coming from such backgrounds,

**At least 85% data completeness on primary outcome measure for trial (repeat compulsory admission within a year):** this was achieved with 69/80 having complete data on the primary outcome (86%). While our target was achieved, this level of missingness is still challenging for reliable inference for a trial: in a future confirmatory trial, we would take additional steps to ensure completeness such as obtaining patient consent to track outcomes via the national Mental Health Services Data Set for those leaving the area.

**At least 60% data completeness for secondary outcomes at 1 year:** this was not achieved, with measures completed for 41/80 participants at a year (51%). Data completeness was however within 20% of our target (41/48: 85% of target). This level of missingness is a major challenge for the trial prohibiting reliable inference regarding secondary measures. We will consider further strategies to address this, for example omitting outcome measures recorded at interview from secondary measures or including only very few.

**At least 75% of intervention participants have developed a crisis plan and have received at least 3 intervention sessions:** this was not reached, with 25/38 (66%) of participants receiving at least three sessions of the intervention and developing a crisis plan. However, the number reaching this minimum target for being defined as having received the intervention was within 20% of our goal (25/28.5, 88%).

Thus, progression criteria were either met, or within 20% of being met: according to our a priori criteria, this suggests we should consider progressing to an application to conduct a full trial after considering improvements that could be made in trial methods and procedures.

The accompanying qualitative study also suggested good acceptability, although the number of participating service users was very small and may well not have been representative (trial participants tended to be reluctant to take part in an interview that was recorded). A number of potential improvements to trial intervention, methods and procedures also emerged from the study, especially the qualitative part. These included ensuring that interventionists (PMHWs) had sufficient time to deliver the intervention, avoiding a situation in which it makes up only a small part of their job plan, and ensuring that they are experienced acute care clinicians who are linked to wards, finding ways to prioritise the crisis planning element of the intervention, and making use of the national Mental Health Services Data Set to avoid loss to follow up due to participants moving catchment areas. Of note also, recruitment to the study began early in 2022, when the impact of COVID-19 on service delivery and on many aspects of community living was still marked: the team’s impression was of continuing disruption to establishing effective community support and meeting practical needs which may have made the intervention more difficult to deliver effectively.

Expert opinion varies regarding the extent to which conclusions should be drawn from findings on potential trial outcomes in feasibility studies such as FINCH which do not have sufficient statistical power to expect a statistically significant finding. The direction of odds ratios in the analysis of primary outcomes favoured the intervention, more so in the sub-group analysis including only people from ethnic minority backgrounds at higher risk of compulsory detention (although any attempt to interpret this observation is impeded not only by small numbers but also by different patterns of diagnosis by ethnic group). The health economic analysis also favoured the intervention, but without reaching statistical significance and with a caveat that inpatient use was already higher at baseline in the control group. These signals suggesting the intervention may prove effective in a larger trial do cohere with the findings of the small body of relevant literature including the original trial in Switzerland (20, 21) and systematic reviews of the literature on approaches to preventing compulsory admissions (12, 13 17). The indication that a greater effect might be anticipated in groups at higher risk of compulsory admission is similar to the finding in a previous randomised controlled trial of advance statements for prevention of compulsory admission, in which larger numbers of Black participants appeared to avoid compulsory admission with an advanced statement, but with insufficient power for investigation of this sub-group (19).

### Limitations

Some limitations to conclusions that can be drawn were inherent in the feasibility study design: statistical power was not sufficient to conduct any definitive analysis of differences in outcomes between groups and the confidence intervals on the estimate of the difference in the candidate primary outcome are wide. The composition of the underlying population from which our sample was recruited is uncertain as recruitment took place intermittently depending on researcher and interventionist availability, and the research team were aware of the target ethnic composition, although in our diverse inpatient population this was achieved without much effort by researchers to recruit from specific groups. Blindness was limited by the small research team and was sometimes broken in extracting the study primary outcome from case notes (although this is possibly less important than for questionnaire ratings as documentation of compulsory detention in clinical records was usually clear). As already described, some progression criteria were not met. Drop out from the follow-up interviews for secondary measures was considerable.

Difficulties in delivering the intervention as planned particularly related to PMHW turnover, with two dropping out and two others going on maternity leave during the study period, while others had only very limited time to deliver the intervention. PMHWs strongly believed that insufficient time had been allocated for the intervention (the time allocated was determined before the study when an application was made for funding to support treatment costs for the study).

The sample of participants willing to take part in a recorded interview for the Phase 2 qualitative study was small. and may have been skewed towards people who had engaged with and had a positive view of the intervention, and analysis of these data was a rapid one carried out with the Theoretical Framework of Acceptability (43) as an a priori approach to organising the analysis.

### Implications of study findings

A degree of reserve is needed in discussing study implications, given its preliminary nature. However, a very important finding is that recruiting compulsorily detained patients in hospital to a trial is feasible: at a time (in the later stages of the COVID-19 pandemic) when many studies have reported difficulty recruiting (44), our recruitment was to time and target, and informal impressions were that both clinicians and inpatients were often very sympathetic to the aims of the study. The literature on how to reduce unnecessary compulsory admissions is very limited in contrast to the importance of this question (8): perceived difficulties in conducting studies with this population is likely to be an important impediment, but our study suggests that research at this stage is feasible (and indeed necessary). The feasibility of recruiting participants from ethnic groups at high risk of detention is also a key finding.

Drop-out rates from both the intervention and the follow-up interviews for secondary outcomes were substantial, and for secondary outcomes would preclude reliable inference in analyses. This is not perhaps a surprise, given that people who have experienced compulsory detention may well have negative views of what mental health services offer and tend not to be well-engaged with services: it was comparable to another recent multicentre trial where a mental health inpatient sample was followed up after discharge (45). However, using a service use-based primary outcome (the obvious one in view of trial aims) helped us achieve relatively high follow-up rates on this outcome. Two thirds of participants received a trial intervention that reached our pre-specified minimum threshold for classifying the intervention as received, formulating a crisis plan and receiving some further check in sessions, again perhaps not an unduly discouraging outcome in this setting. Those participants (only a small minority) with whom we were able to conduct a qualitative interview found the intervention acceptable, as did clinicians delivering it except for lack of time, a potentially remediable difficulty.

Thus findings suggest that this research direction is worth pursuing especially as results cohere with previous evidence and there is a pressing need to understand how to reduce numbers of coercive, distressing and expensive compulsory inpatient admissions.

Inspection of findings on the primary outcome suggests a possibility that the intervention may be more successful among people from ethnic backgrounds at high risk of compulsory admission, which might be explained in terms of this group currently being under-served and too often regarded as unlikely to engage with psychosocial interventions that may in reality be helpful if offered. Calculations of the number needed to treat suggest a full trial may also be more feasible in this group, with an estimate that the intervention needs to be offered to 9 people for a compulsory detention to be prevented. Delivering the intervention for this degree of benefit is likely to be worthwhile given the relatively low cost of the intervention and the great benefits associated with avoiding admission, a goal that there are currently no other evidence-based strategies to achieve. However, a large sample size is likely to be needed for good power to detect an effect in a full trial, with attention also to the potential improvements to trial and intervention processes suggested by this feasibility study.

Approaches worth considering for a full trial include assessing only routinely recorded outcomes such as compulsory hospitalisation only, and increasing PMHW time and flexibility for delivering the intervention. Further consideration of who delivers the intervention may also be helpful: following our Swiss model, our initial preference was for clinical psychologists to deliver the intervention. However, we experienced some difficulties in recruiting and retaining them, leading us to deploy other professionals: our impression was that experience in working flexibly with a population with relatively severe difficulties and ambivalence towards mental health services, and in delivering structured interventions was more important than type of professional. A recently published study from France successfully engaged peer support workers to deliver an intervention based on advance statements (22), and a previous study from our group found a peer support worker-delivered intervention to be effective following discharge from crisis resolution teams (46). We considered intervention delivery by peer support workers in the current study, but eventually decided to follow previous studies, especially our Swiss model, in employing qualified professionals. Our concerns included that working with a high level of independence with a patient group potentially at high risk of relapse and of losing capacity might exceed reasonable expectations regarding peer support worker roles (47). However, peer support workers increasingly work in a wide range of mental health settings and may have particular strengths in engaging a population who tend to be disillusioned with mental health services, so that their involvement in interventions to prevent compulsory re-detention seems well worth considering further.

In relation to service planning, supported self-management (26) and crisis planning (12) already have substantial supporting evidence: our study suggests that delivering such interventions around the time of discharge is feasible and acceptable for at least some inpatients. Even in a system characterised by brief admissions and a tendency for patients on wards to be acutely unwell, obtaining the informed consent of inpatients to receive structured interventions proved to be feasible and may present a good opportunity to initiate therapeutic relationships and treatment engagement (22), fostering hope that repeat compulsory admission can be avoided and meaningful recovery achieved in future.

## Data Availability

All data produced in the present study are available upon reasonable request to the authors

## Acknowledgements

The authors would like to thank the many people who contributed to this study, including research delivery teams and clinicians in each of the participating Trusts and of course the 80 inpatients who agreed to be randomised to test the study intervention.

## Funding

This report describes independent research funded by the National Institute of Health and Care Research Policy Research Programme (Ref: NIHR20173). The views expressed are those of the authors and not necessarily of the NIHR or the Department of Health and Social Care Research. The funders have no role in the design of this study and will not have any role during its execution, data analyses, and interpretation, but are expected to provide assistance to any enquiry, audit, or investigation related to the funded work.

The authors have no conflicts of interest to declare.

Availability of data and draft intervention manual: On request from lead author

## Online Appendix A Intervention and research activities in the Finch trial as reported in current paper

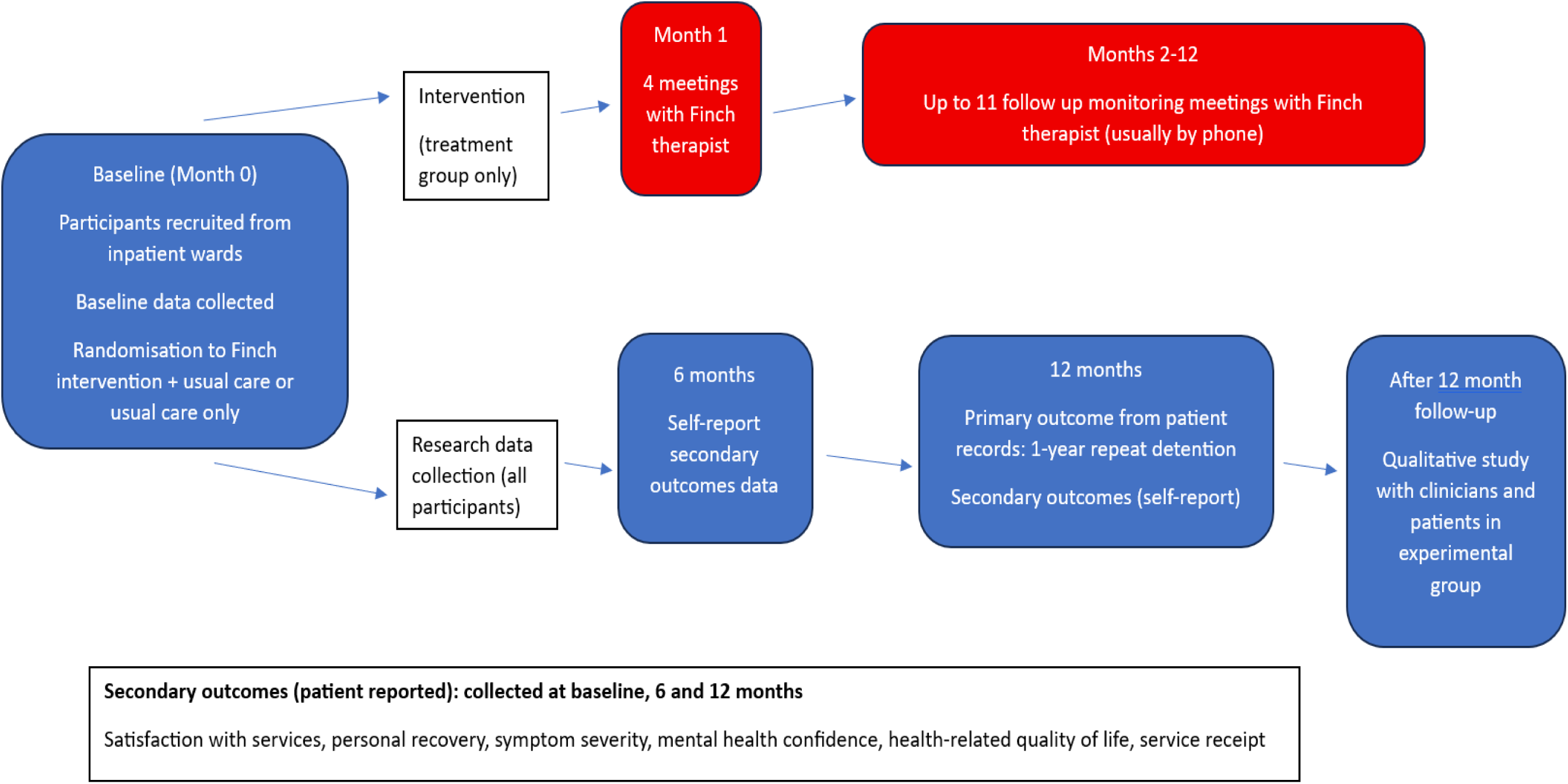

## Online Appendix B Phase 2 Interview Topic Guides

### Interview Topic Guide for interviews with *Personal Mental Health Workers (PMHWs)* for Phase Two Qualitative Study: to investigate experiences of receiving and delivering the intervention

#### Introduction

We are looking to understand your perspective on the crisis-planning intervention you have been delivering. We are speaking to the personal mental health workers who have been delivering the intervention over the past six-nine months. We’d like to discuss how you have found delivering the intervention, understand your view on the impact it may have had on service users, and your ideas for how it could be improved in the future.

1. In general, could you tell me about your experience of delivering the crisis-planning intervention over the past six-nine months? Prompt: for: attitude to the intervention (both positive and negative aspects), both initial four therapy sessions and monthly check-ins.
2. Which parts/components of the intervention, if any, do you think have been more helpful for the people you’ve been working with? Prompts: These are some of the components that may or may not have been helpful to your patients: having the first four sessions take place on the ward, developing a narrative about how the patient came to be detained on the ward, developing a formulation about why the sectioning occurred, developing a crisis plan, advanced statement and setting recovery goals, message to future self, monthly follow-up sessions, reviewing crisis plans, advanced statement or recovery goals.
3. Overall, would you say that the intervention is having a positive, negative or neutral influence on people’s recovery? Why do you think this?
4. Which components of the intervention, if any, do you think have been less helpful? Were there any particular parts of the intervention that you think the patient found less helpful? Prompt for: difficulties with discussing sensitive matters and traumatic experiences, difficulties with relationship with professionals, specific elements such as crisis plans and advance statements, practical challenges such as setting up appointments.
5. What was your relationship(s) like with your patients(s)? Prompt for: What helped to build a relationship with your patients? What were the barriers or challenges to building a relationship? Do you think and interpersonal or demographic differences between you and the patient impacted on the relationship?
6. Did you notice any changes in your patient(s) as a result in taking part in the intervention? Prompt: what changes did you notice?
7. Were there aspects of the intervention that were challenging or difficult to put into practice? Prompts: participant factors such as their current mental state, location of sessions, completing formulation, crisis plans, advance statement, setting recovery goals, keeping in contact with participants.
8. Did you have any patients drop out or disengage? What led them to dropping out or disengaging? Prompt: Do you have any ideas about what might prevent people from dropping out or disengaging with therapy?
9. What would you change about the intervention to make it more helpful for people? Prompt for: specific aspects of the intervention content; how check-in calls could have been improved; any new ideas for useful content to be included; views on the intervention length (four sessions and 12 x monthly check-ins); views on the therapeutic alliance between service user and clinician and ideas for improvement
10. What ideas do you have for making the intervention easier for clinicians to deliver? Prompt for: ideas for the four initial sessions, ideas for the check-in sessions (views on frequency, format and length of check-ins), aspects of the intervention that were more or less feasible from the clinician’s perspective, any trade-offs or extra burdens delivering the intervention led to for clinicians
11. How do you think the intervention has fitted in with other parts of the care the participants have been receiving? Prompt for: whether communicating with other staff involved in care, whether it fits with overall care plan, whether it duplicates or adds to this.
12. What was your experience of the supervision (monthly group supervision provision)? Prompt: How do you think this could be improved? Is there anything else that could have been put in place to make you feel more supported?
13. What was your experience of the training (i.e two half days and one full day online training? Prompt: How do you think the training could be improved? Is there anything else you would liked to have seen in the training?
14. Is there anything else that you would like to share that we haven’t covered? We have now come to the end of the interview. Do you have any questions? Thank you for your time.

### Interview Topic Guide for interviews with Service users for Phase Two Qualitative Study: to investigate experiences of receiving and delivering the intervention

#### Introduction

We would like to know how you have found the new type of support which involved crisis-planning that you have been receiving as part of the FINCH study that you kindly agreed to take part in. This new type of support involved you receiving additional help from [name of person who delivered intervention to the person] a personal mental health worker who you met with for four weekly one to one sessions on the ward, or maybe just after you were discharged, and then eleven monthly calls online of over the phone.

The focus was on developing Crisis Plans, an advance statement and setting recovery goals. In this interview we will discuss your experiences since being discharged from hospital. We can stop the interview at any time if you find discussing any of the topics difficult or upsetting and please ask if anything is not clear.

#### Do you have any questions you’d like to ask us before we start?

1. Can you tell me about your experiences of working with [name of person delivering the intervention] to receive the additional support I mentioned just now? You don’t need to go into too much detail, just a summary for now. Prompts: What did you cover/discuss? What was the focus of the sessions?
2. Which parts of this new type of support, if any, have been most helpful to you? Prompt for: were the discussions helpful, any helpful aspects of practical arrangements, and content of sessions, including monthly check ins, developing an understanding of what happened in run-up being “sectioned”, development of crisis plan, advance statement, recovery goals, “message to future self”/
3. Which parts of the new type of support, if any, have been less helpful or challenging for you? Prompt for which were challenging components of sessions, which were less helpful, which were burdensome or not practical
4. What was your relationship like with [insert clinician’s name]? OR how did you get on with [insert clinician’s name]? Prompt: Were you made to feel comfortable/safe/at ease/respected. Could you discuss race, ethnicity or culture?
5. How did the intervention fit into other forms of support you received, for example, from other mental health staff or clinicians? Prompt for: fit with work with Care co-ordinator, other community mental health team staff.
6. Have you noticed any changes as a result of taking part in the new type of support? What changes have you noticed? Prompt for change in understanding of what may lead to relapse or detention, new strategies to use if becoming unwell, any other changes that might result from this work.
7. What were the main things you got out of the new type of support? What were the key take home messages?
8. Which factors in your life have had the biggest influence on being able to get on with your life following your discharge from hospital and/or on your mental health and staying well? Prompt for: positive and negative external and internal influences on recovery and well-being; which factors were more influential and which factors were less influential (factors could include: social support, employment, physical health needs, housing, financial support etc.)
9. Has participating in this new type of support influenced your recovery in any way? Why do you think this is? Prompts: effects on self-confidence, positive and negative effects on recovery after hospital, any effects on coping in crisis situations, reasons why new type of support helpful overall or not.
10. What would you change about the new type of support to make it more helpful Prompt for changes in: format (four main initial sessions, then monthly check-ins), developing a crisis plan, developing an advance statement, setting recovery goals, relationship with clinician, any other content that would have liked)
11. Would you recommend this new type of support to someone else in the same position? Please explain your answer.
12. Are there any questions you would like to have been asked, that we didn’t ask?

## Online Appendix C Findings for proposed primary outcome for whole sample and for groups at higher and lower risk of compulsory detention

**Table C1:** Odds Ratio of Not Being Detained – whole sample.

|  | Allocation |  |  |  | Odds Ratio | (95% CI) | p-Value |
| --- | --- | --- | --- | --- | --- | --- | --- |
|  | Control [N = 42] |  | Intervention [N = 38] |  |  |  |  |
|  | N | (%) | N | (%) |  |  |  |
| Not Detained at 12 Months | 23 | (67.6) | 26 | (74.3) | 1.378 | (0.480 to 3.955) | 0.551 |
| Adjusted for Risk Level |  |  |  |  |  |  |  |

Risk Difference
|  | Allocation |  |  |  | Risk<br>Difference | (95% CI) |
| --- | --- | --- | --- | --- | --- | --- |
|  | Control [N =<br>42] |  | Intervention [N =<br>38] |  |  |  |
|  | N | (%) | N | (%) |  |  |
| Not Detained at 12<br>Months | 23 | (67.6) | 26 | (74.3) | 0.065 | (-0.147 to<br>0.276) |
| Adjusted for Risk Level |  |  |  |  |  |  |

Risk Ratio
|  | Allocation |  |  |  | Risk Ratio | (95% CI) |
| --- | --- | --- | --- | --- | --- | --- |
|  | Control [N = 42] |  | Intervention [N = 38] |  |  |  |
|  | N | (%) | N | (%) |  |  |
| Not Detained at 12 Months | 23 | (67.6) | 26 | (74.3) | 1.095 | (0.766 to 1.424) |
| Adjusted for Risk Level |  |  |  |  |  |  |

Number Needed to Treat
The Number Needed to Treat (NNT) = 16

**Table C2.** Restricted to High-Risk Group (people from ethnic backgrounds at higher risk of compulsory detention)

|  | Allocation |  |  |  | Odds Ratio | (95% CI) |
| --- | --- | --- | --- | --- | --- | --- |
|  | Control [N = 21] |  | Intervention [N = 19] |  |  |  |
|  | N | (%) | N | (%) |  |  |
| Not Detained at 12 Months | 10 | (58.8) | 12 | (70.6) | 1.680 | (0.405 to 6.962) |

Risk Difference
|  | Allocation |  |  |  | Risk<br>Difference | (95% CI) |
| --- | --- | --- | --- | --- | --- | --- |
|  | Control [N =<br>21] |  | Intervention [N =<br>19] |  |  |  |
|  | N | (%) | N | (%) |  |  |
| Not Detained at 12<br>Months | 10 | (58.8) | 12 | (70.6) | 0.118 | (-0.201 to<br>0.436) |

Risk Ratio
|  | Allocation |  |  |  | Risk Ratio | (95% CI) |
| --- | --- | --- | --- | --- | --- | --- |
|  | Control [N = 21] |  | Intervention [N = 19] |  |  |  |
|  | N | (%) | N | (%) |  |  |
| Not Detained at 12 Months | 10 | (58.8) | 12 | (70.6) | 1.200 | (0.597 to 1.803) |

The Number Needed to Treat (NNT) = 9

## Appendix D Proposed trial secondary measures

Table D1 shows findings for secondary measures collected at interview. No statistical analyses beyond descriptive statistics were planned for these measures given the status of the trial as a feasibility study. Table D2 shows BPRS scores item by item. A total score was not computed for BPRS as the completion of many follow-up interviews by phone meant that not all the observer-rated items on the BPRS could be rated, leaving very small numbers for total scores at follow-up. We therefore report on the BPRS only as mean scores for each item at each time point.

**Table D1:** Scores for secondary outcome measures at all time points.

|  | Control Group n=42 |  | Intervention=38 |  |
| --- | --- | --- | --- | --- |
|  | mean | (sd) | mean | (sd) |
| Client Satisfaction Questionnaire Baseline | 20.35<br>n=40 | (6.29) | 19.92<br>n=38 | (6.31) |
| Client Satisfaction Questionnaire 6 months | 20.00<br>n=26 | (6.45) | 24.95<br>n=20 | (5.22) |
| Client Satisfaction Questionnaire 12 Months | 20.43<br>n=24 | (8.03) | 24.75<br>n=17 | (5.13) |
| Mental Health Confidence scale Baseline | 63.4<br>n=40 | (19.15) | 61.55<br>n=38 | (21) |
| Mental Health Confidence scale 6 months | 60.92<br>n=26 | (19.10) | 58.33<br>n=20 | (11.17) |
| Mental Health Confidence scale 12 months | 61.17<br>n=24 | (17.39) | 58.06<br>n=17 | (18.35) |
| The Process of Recovery Questionnaire Baseline | 40.51<br>n=40 | (13.84) | 41.49<br>n=38 | (12.01) |
| The Process of Recovery Questionnaire 6 months | 34.58<br>n=26 | (14.56) | 36.59<br>n=20 | (9.92) |
| The Process of Recovery Questionnaire 12 months | 37.12<br>n=24 | (14.22) | 38.35<br>n=17 | (14.42) |

### Appendix D2 Means for BPRS items at each timepoint where available

| BPRS Item | Baseline |  |  | Six months |  |  | Twelve months |  |  |
| --- | --- | --- | --- | --- | --- | --- | --- | --- | --- |
|  | Intervention | Control | Total | Intervention | Control | Total | Intervention | Control | Total |
|  | n=38 | n=40 | n=78 | n= 17 | n=26 | n=43 | n=11 | n = 22 | n=33 |
| 1. Somatic concern | 3.05 | 2.45 | 2.82 | 2.17 | 2.42 | 2.32 | 1.82 | 2.91 | 2.54 |
| 2. Anxiety | 3.97 | 3.55 | 3.75 | 3.41 | 3.38 | 3.39 | 4.00 | 3.59 | 3.72 |
| 3. Depression | 3.73 | 3.72 | 3.73 | 3.47 | 2.96 | 3.16 | 3.64 | 3.05 | 3.24 |
| 4. Suicidality | 2.31 | 2.25 | 2.30 | 1.94 | 1.88 | 1.90 | 2.27 | 1.86 | 2.00 |
| 5. Guilt | 2.63 | 2.67 | 2.65 | 2.88 | 2.42 | 2.60 | 2.36 | 2.41 | 2.39 |
| 6. Hostility | 2.44 | 2.55 | 2.34 | 1.58 | 1.42 | 1.48 | 1.73 | 1.82 | 1.78 |
| 7. Elated Mood | 2.02 | 1.62 | 1.82 | 1.82 | 1.15 | 1.41 | 1.00 | 1.36 | 1.24 |
| 8. Grandiosity | 2.31 | 2.25 | 2.28 | 1.29 | 1.07 | 1.16 | 1.27 | 1.23 | 1.24 |
| 9. Suspiciousness | 3.02 | 2.25 | 2.62 | 1.88 | 2.07 | 2 | 2.09 | 2.05 | 2.06 |
| 10. Hallucinations | 2.23 | 2.07 | 2.15 | 1.76 | 2.03 | 1.93 | 1.00 | 2.00 | 1.66 |
| 11. Unusual thought content | 2.44 | 2.12 | 2.28 | 1.47 | 1.34 | 1.39 | 1.09 | 1.36 | 1.27 |
|  |  |  |  | n=7 | n=12 | n=19 | n=0 | n=5 | n=5 |
| 12. Bizarre behaviour | 1.92 | 1.77 | 1.86 | 1.14 | 1.66 | 1.47 | 0 | 1 | 1 |
| 13. Self-neglect | 1.6 | 2 | 1.80 | 2 | 2 | 2 | 0 | 1.8 | 1.8 |
| 14. Disorientation | 1.15 | 1.32 | 1.24 | 1.14 | 1.08 | 1.10 | 0 | 1 | 1 |
| 15. Conceptual disorganisation | 1.42 | 1.52 | 1.47 | 1.14 | 1 | 1.05 | 0 | 1 | 1 |
| 16. Blunted affect | 1.81 | 2.7 | 2.26 | 1.71 | 1.41 | 1.52 | 0 | 1 | 1 |
| 17. Emotional withdrawal | 1.34 | 1.75 | 1.55 | 1.28 | 1.33 | 1.31 | 0 | 1 | 1 |
| 18. Motor retardation | 1.5 | 1.77 | 1.64 | 1.71 | 1.41 | 1.52 | 0 | 1 | 1 |
| 19. Tension | 1.57 | 1.85 | 1.71 | 1.14 | 1.58 | 1.42 | 0 | 1 | 1 |
| 20. Uncooperativeness | 1.23 | 1.6 | 1.42 | 1.14 | 1.08 | 1.10 | 0 | 1 | 1 |
| 21. Excitement | 1.47 | 1.67 | 1.57 | 1.14 | 1 | 1.05 | 0 | 1 | 1 |
| 22. Distractibility | 1.55 | 1.92 | 1.74 | 1.28 | 1.16 | 1.21 | 0 | 1 | 1 |
| 23. Motor hyperactivity | 1.42 | 1.52 | 1.47 | 1.14 | 1.33 | 1.26 | 0 | 1.2 | 1.2 |
| 24. Mannerisms and posturing | 1.13 | 1.175 | 1.15 | 1 | 1.25 | 1.15 | 0 | 1 | 1 |

Large amounts of missing data on some items at follow-up reflects interviews that were carried out by telephone based on participants' preferences, so that items based on observation could not be assessed.

## Appendix E Health economic analysis

The cost of the intervention was based on the average unit cost for a psychologist, mental health nurse, and occupational therapist taken from the NHS Refence Costs for 2021/2. The cost per session was £120. It was assumed that the length of monthly catch-up meetings lasted for 75% of a usual contact. Other service use was measured using the Client Service Receipt Inventory (CSRI) (41) and covered the six-month periods prior to baseline and 6- and 12-month follow-up. Services included primary and secondary care as well as social care services. Unit costs were obtained from the annual compendium published by the Universities of York and Kent and the NHS Reference Costs. The number and percentage of participants using services at each time point are reported, along with the mean and standard deviation (sd) number of contacts for those with at least one contact, and the mean (sd) cost for all participants (i.e. including those with a zero cost for that service). Mean total costs for the combined follow-up period are compared with bootstrapped 95% confidence intervals generated around the cost difference between the two groups.

The EQ-5D-5L (39) was used to measure health-related quality of life for use as an outcome measure in the economic evaluation. This consists of five components: mobility, self-care, usual activities, pain and discomfort, and anxiety and depression. Each are rated with an integer between 1 (no problem) and 5 (extreme problems). However, it was only used at 12-month follow-up and so quality-adjusted life years (QALYs) could not be computed. Utility values were though derived from an algorithm and compared between the two groups.

Total costs could be computed from the CSRI for 40 (95.2%) usual care and 36 (94.7%) intervention participants at baseline, 36 (85.7%) usual care and 37 (97.4%) intervention participants at 6-month follow-up, and 29 (69.0%) usual care and 34 (89.5%) intervention participants at 12-month follow-up.

In the six months prior to baseline all participants spent time as inpatients (Table E1 below). The mean duration of index admission during which recruitment took place for those in the intervention group for whom we have data was 86 days (n=36), and 108 days for those in the control group (n=37). Almost all were also inpatients for some of the period before the 6-month follow-up, but in the six months prior to the 12-month follow-up there were fewer in the intervention group who had inpatient stays. Number of participants detained under MHA within six months of baseline in the intervention group (n=36) was 7 (19.4%), and 9 (25/0%) in control group (n=36). In Use of GPs was slightly more likely for the intervention group prior to baseline, but prior to the 12-month follow-up this had switched with more of the usual care group seeing a GP. Contacts with psychiatrists, mental health nurses, and other mental health workers were common for both groups in each period but without major differences between groups. Prior to the 12-month follow-up, the usual care group were more likely to have had contact with other doctors. Day care services were used by far fewer participants.

The average number of inpatient days for those with admissions (i.e. excluding those with zero use) was substantially higher at baseline and in the period up to 6-month follow-up for the usual care group (Table E2 below). In the period before the 12-month follow-up the average number of days was higher for the intervention group (but from Table 5 we saw the proportion with stays was greater for the usual care group). The mean number of contacts by users of other services was highly variable due to many having relatively few users.

The mean (SD) intervention cost was £583 (£404) for the intervention group, with a range of £0 to £1385. Other care costs were dominated by inpatient costs (Table E3 below). The were noticeably higher for the usual care group at each time point. After inpatient care, the highest costs were for crisis team contacts, psychiatrists, mental health nurses, and other mental health workers. The total mean (sd) cost over the combined follow-up period, including the intervention, was £41,840 (£29,068) for the usual care group and £35,962 (£38,020) for the intervention group. The mean incremental cost was −£5872 (i.e. lower for the intervention group) with a bootstrapped 95% confidence interval of −£22,204 to £9781, showing the difference not to be statistically significant.

Th EQ-5D-5L utility score at 12-month follow up was available for 24 (57.1%) usual care and 17 (44.7%) intervention participants. The mean (sd) score was 0.667 (0.246) for the usual care group and 0.675 (0.318) for the intervention group. The difference of 0.008 was not statistically significant (95% CI, −0.170 to 0.186).

ReQoL utility scores could be calculated for 37 (97.3%) intervention group and 39 (92.8%) control group participants at baseline, 17 (44.7%) intervention group and 26 (61.9%) control group participants at 6-month follow-up, and 12 (31.6%) intervention group and 22 (52.4%) control group participants at 12-month follow-up. The mean (SD) utility scores at baseline were 0.718 (0.236) for the intervention group and 0.804 (0.185) for the control group. At 6-month follow-up the scores were 0.804 (0.143) for the intervention group and 0.739 (0.270) for the control group. The intervention group had a score that was 0.065 higher (95% CI, −0.079 to 0.210). At 12-month follow-up the scores were 0.760 (0.268) for the intervention group and 0.770 (0.231) for the control group. The intervention group had a score that was 0.010 lower (95% CI, −0.189 to 0.169).

**Table E1.**
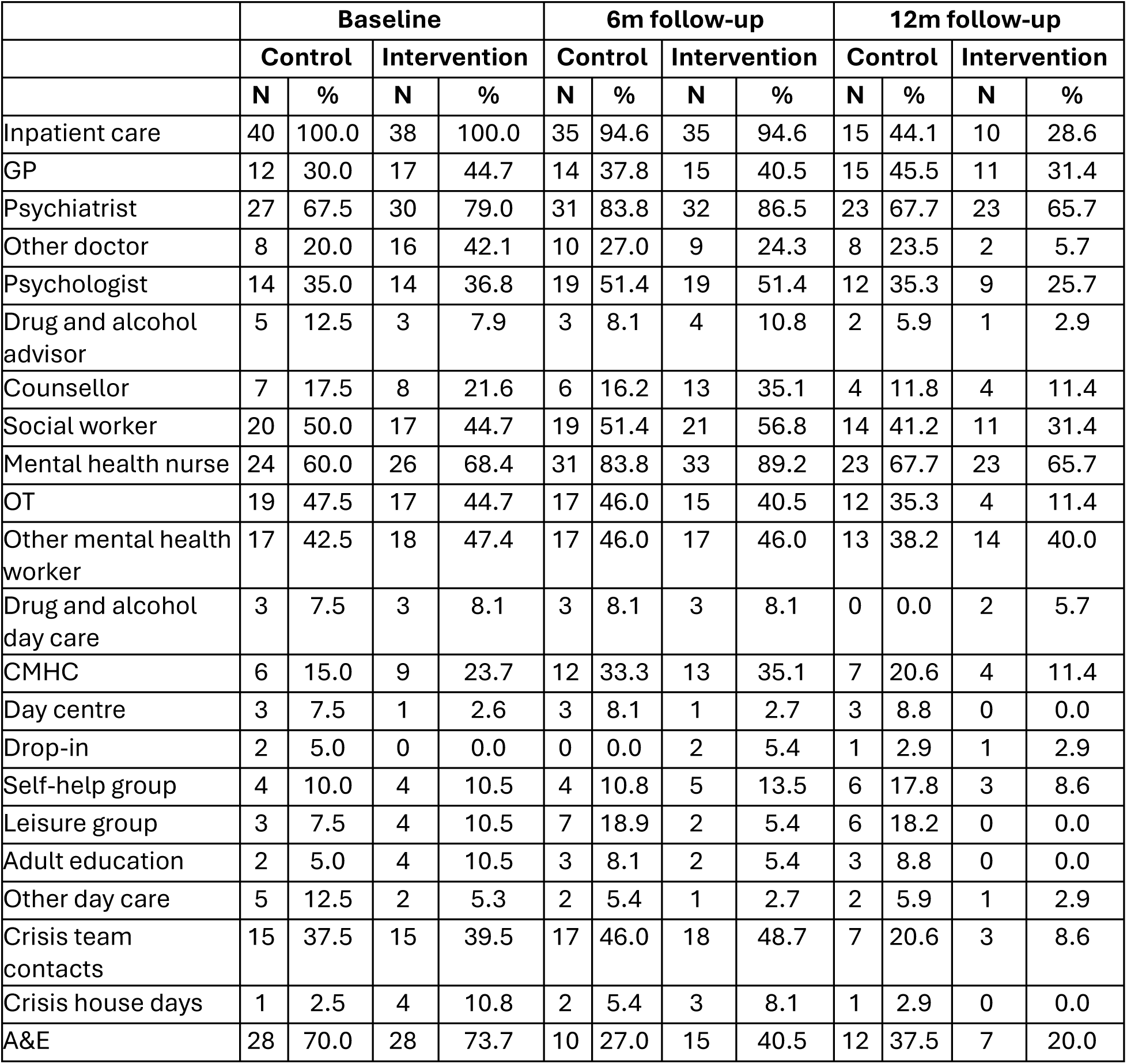
Number and % using services in 6 months prior to baseline and each follow-up.

|  | Baseline |  |  |  | 6m follow-up |  |  |  | 12m follow-up |  |  |  |
| --- | --- | --- | --- | --- | --- | --- | --- | --- | --- | --- | --- | --- |
|  | Control |  | Intervention |  | Control |  | Intervention |  | Control |  | Intervention |  |
|  | N | % | N | % | N | % | N | % | N | % | N | % |
| Inpatient care | 40 | 100.0 | 38 | 100.0 | 35 | 94.6 | 35 | 94.6 | 15 | 44.1 | 10 | 28.6 |
| GP | 12 | 30.0 | 17 | 44.7 | 14 | 37.8 | 15 | 40.5 | 15 | 45.5 | 11 | 31.4 |
| Psychiatrist | 27 | 67.5 | 30 | 79.0 | 31 | 83.8 | 32 | 86.5 | 23 | 67.7 | 23 | 65.7 |
| Other doctor | 8 | 20.0 | 16 | 42.1 | 10 | 27.0 | 9 | 24.3 | 8 | 23.5 | 2 | 5.7 |
| Psychologist | 14 | 35.0 | 14 | 36.8 | 19 | 51.4 | 19 | 51.4 | 12 | 35.3 | 9 | 25.7 |
| Drug and alcohol advisor | 5 | 12.5 | 3 | 7.9 | 3 | 8.1 | 4 | 10.8 | 2 | 5.9 | 1 | 2.9 |
| Counsellor | 7 | 17.5 | 8 | 21.6 | 6 | 16.2 | 13 | 35.1 | 4 | 11.8 | 4 | 11.4 |
| Social worker | 20 | 50.0 | 17 | 44.7 | 19 | 51.4 | 21 | 56.8 | 14 | 41.2 | 11 | 31.4 |
| Mental health nurse | 24 | 60.0 | 26 | 68.4 | 31 | 83.8 | 33 | 89.2 | 23 | 67.7 | 23 | 65.7 |
| OT | 19 | 47.5 | 17 | 44.7 | 17 | 46.0 | 15 | 40.5 | 12 | 35.3 | 4 | 11.4 |
| Other mental health worker | 17 | 42.5 | 18 | 47.4 | 17 | 46.0 | 17 | 46.0 | 13 | 38.2 | 14 | 40.0 |
| Drug and alcohol day care | 3 | 7.5 | 3 | 8.1 | 3 | 8.1 | 3 | 8.1 | 0 | 0.0 | 2 | 5.7 |
| CMHC | 6 | 15.0 | 9 | 23.7 | 12 | 33.3 | 13 | 35.1 | 7 | 20.6 | 4 | 11.4 |
| Day centre | 3 | 7.5 | 1 | 2.6 | 3 | 8.1 | 1 | 2.7 | 3 | 8.8 | 0 | 0.0 |
| Drop-in | 2 | 5.0 | 0 | 0.0 | 0 | 0.0 | 2 | 5.4 | 1 | 2.9 | 1 | 2.9 |
| Self-help group | 4 | 10.0 | 4 | 10.5 | 4 | 10.8 | 5 | 13.5 | 6 | 17.8 | 3 | 8.6 |
| Leisure group | 3 | 7.5 | 4 | 10.5 | 7 | 18.9 | 2 | 5.4 | 6 | 18.2 | 0 | 0.0 |
| Adult education | 2 | 5.0 | 4 | 10.5 | 3 | 8.1 | 2 | 5.4 | 3 | 8.8 | 0 | 0.0 |
| Other day care | 5 | 12.5 | 2 | 5.3 | 2 | 5.4 | 1 | 2.7 | 2 | 5.9 | 1 | 2.9 |
| Crisis team contacts | 15 | 37.5 | 15 | 39.5 | 17 | 46.0 | 18 | 48.7 | 7 | 20.6 | 3 | 8.6 |
| Crisis house days | 1 | 2.5 | 4 | 10.8 | 2 | 5.4 | 3 | 8.1 | 1 | 2.9 | 0 | 0.0 |
| A&E | 28 | 70.0 | 28 | 73.7 | 10 | 27.0 | 15 | 40.5 | 12 | 37.5 | 7 | 20.0 |

**Table E2.** Mean and SD contacts for those using services in 6 months prior to baseline and each follow-up.

|  | Baseline |  |  |  | 6m follow-up |  |  |  | 12m follow-up |  |  |  |
| --- | --- | --- | --- | --- | --- | --- | --- | --- | --- | --- | --- | --- |
|  | Control |  | Intervention |  | Control |  | Intervention |  | Control |  | Intervention |  |
|  | Mean | SD | Mean | SD | Mean | SD | Mean | SD | Mean | SD | Mean | SD |
| Inpatient days | 52.8 | 43.0 | 40.0 | 39.0 | 59.6 | 55.0 | 44.7 | 50.0 | 73.1 | 55.1 | 93.9 | 73.4 |
| GP | 3.5 | 3.8 | 2.8 | 3.5 | 3.9 | 3.2 | 1.5 | 0.7 | 2.2 | 1.3 | 1.9 | 1.2 |
| Psychiatrist | 5.2 | 3.4 | 4.6 | 4.0 | 5.6 | 5.3 | 5.8 | 5.3 | 4.9 | 5.1 | 4.5 | 5.6 |
| Other doctor | 1.9 | 0.8 | 2.5 | 2.1 | 2.4 | 2.4 | 2.2 | 1.5 | 4.1 | 2.9 | 3.5 | 3.5 |
| Psychologist | 3.4 | 4.6 | 3.6 | 3.8 | 4.7 | 4.7 | 5.6 | 6.7 | 5.3 | 6.4 | 7.3 | 5.8 |
| Drug and alcohol advisor | 19.6 | 39.4 | 6.3 | 9.2 | 2.3 | 2.3 | 5.8 | 6.9 | 1.5 | 0.8 | 1.0 | - |
| Counsellor | 3.2 | 3.6 | 5.6 | 8.3 | 2.6 | 2.5 | 9.3 | 12.0 | 8.0 | 9.1 | 10.0 | 9.5 |
| Social worker | 3.6 | 3.8 | 3.8 | 4.8 | 3.5 | 3.6 | 4.1 | 3.5 | 4.8 | 6.4 | 4.1 | 4.3 |
| Mental health nurse | 7.2 | 8.4 | 9.1 | 10.0 | 18.4 | 20.9 | 19.1 | 28.9 | 17.9 | 19.2 | 8.7 | 10.8 |
| OT | 6.2 | 6.9 | 4.7 | 3.9 | 7.3 | 6.5 | 8.7 | 9.8 | 4.8 | 4.7 | 19.8 | 19.8 |
| Other mental health worker | 12.2 | 14.2 | 11.2 | 8.6 | 15.4 | 14.6 | 15.9 | 17.7 | 10.8 | 8.9 | 23.1 | 47.3 |
| Drug and alcohol day care | 1.0 | 0.0 | 1.0 | 0.0 | 5.7 | 5.5 | 2.7 | 1.5 | - | - | 2.0 | 0.0 |
| CMHC | 6.3 | 9.4 | 3.4 | 2.4 | 4.6 | 5.0 | 7.5 | 6.2 | 14.0 | 11.2 | 11.5 | 9.0 |
| Day centre | 1.0 | 0.0 | 2.0 | - | 2.7 | 2.9 | 2.0 | - | 6.0 | 4.0 | - | - |
| Drop-in | 2.0 | 1.4 | - | - | - | - | 3.5 | 2.1 | 10.0 | - | 1.0 | - |
| Self-help group | 2.8 | 3.5 | 40.5 | 77.0 | 4.3 | 1.5 | 4.6 | 4.3 | 16.7 | 21.0 | 9.3 | 12.7 |
| Leisure group | 9.0 | 13.0 | 35.8 | 62.9 | 12.1 | 13.3 | 24.5 | 33.2 | 15.0 | 12.8 | - | - |
| Adult education | 2.0 | 0.0 | 3.0 | 3.4 | 1.0 | 0.0 | 24.5 | 33.2 | 17.7 | 15.9 | - | - |
| Other day care | 24.0 | 27.8 | 15.0 | 15.6 | 1.5 | 0.7 | 2.0 | - | 32.3 | 5.3 | 21.0 | - |
| Crisis team contacts | 10.5 | 11.3 | 14.7 | 28.0 | 35.2 | 28.0 | 24.5 | 21.5 | 22.9 | 13.5 | 15.3 | 11.6 |
| Crisis house days | 14.0 | 0.0 | 6.0 | 3.4 | 12.0 | 2.8 | 5.5 | 4.0 | 14.0 | 0.0 | - | - |
| A&E | 2.1 | 1.5 | 2.3 | 2.0 | 1.8 | 1.3 | 1.7 | 1.3 | 2.3 | 1.9 | 2.1 | 1.5 |

**Table E3.**
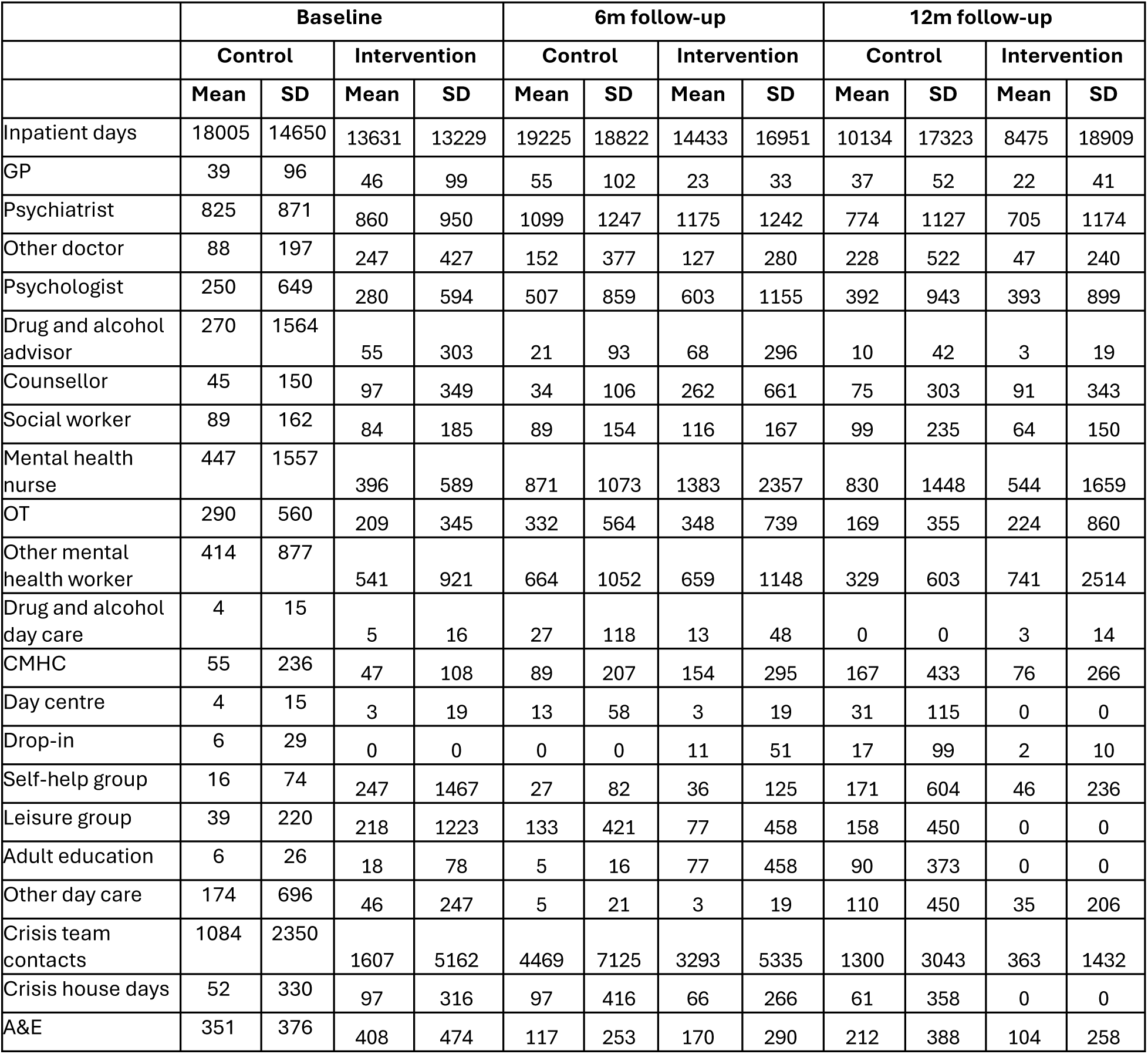
Mean and SD service cost in 6 months prior to baseline and each follow-up (2021/2 £s).

## Footnotes

1 White Other is a category used in UK official data collection to include all White groups other than White British and White Irish, the largest groups being from other European countries, North America and Australasia.

## Notes

### Competing Interest Statement

The authors have declared no competing interest.

### Clinical Trial

ISRCTN11627644

### Clinical Protocols

https://pilotfeasibilitystudies.biomedcentral.com/articles/10.1186/s40814-024-01453-z#Fun

### Author Declarations

Full Health Research Authority (HRA) and NHS Research Ethics Committee (REC) approval was granted by the London-Bromley Research Ethics Committee (IRAS: 300671; Protocol number: 143180; REC reference: 21/LO/0734). The trial sponsor was the Joint Research Office for UCL and UCLH (Ref: 143180).

### Summary of Updates

I have added a co-author who was omitted in error - Valerie Christina White. Apologies for this.

## References

1. Sheridan Rains L, Zenina T, Dias MC, Jones R, Jeffreys S, Branthonne-Foster S….. & Johnson, S. Variations in patterns of involuntary hospitalisation and in legal frameworks: an international comparative study. Lancet Psychiatry. 2019;6(5):403–17.

2. Barnett P, Mackay E, Matthews H, Gate R, Greenwood H, Ariyo K ….& Smith, S. Ethnic variations in compulsory detention under the Mental Health Act: a systematic review and meta-analysis of international data. Lancet Psychiatry. 2019;6(4):305–17.

3. NHS Digital Mental Health Act Statistics, Annual Figures England 2023-2024. Mental Health Act Statistics, Annual Figures - NHS England Digital. Accessed 26/02/2024

4. Akther SF, Molyneaux E, Stuart R, Johnson S, Simpson A, Oram S. Patients’ experiences of assessment and detention under mental health legislation: systematic review and qualitative meta-synthesis. BJPsych Open. 2019;5(3):e37.

5. Stuart R, Akther SF, Machin K, Persaud K, Simpson A, Johnson S & Oram S. Carers’ experiences of involuntary admission under mental health legislation: systematic review and qualitative meta-synthesis. BJPsych Open. 2020;6(2):e19.

6. Bartl, G., Stuart, R., Ahmed, N., Saunders, K., Loizou, S……& Lloyd-Evans, B. A qualitative meta-synthesis of service users’ and carers’ experiences of assessment and involuntary hospital admissions under mental health legislations: a five-year update. BMC Psychiatry 24, 476 (2024).

7. Priebe S, Katsakou C, Yeeles K, Amos T, Morriss R, Wang D, Wykes T. Predictors of clinical and social outcomes following involuntary hospital admission: a prospective observational study. Eur Arch Psychiatry Clin Neurosci. 2011;261(5):377–86.

8. Bone JK, McCloud T, Scott HR, Machin K, Markham S, Persaud K, et al. Psychosocial Interventions to Reduce Compulsory Psychiatric Admissions: A Rapid Evidence Synthesis. EClinicalMedicine. 2019;10:58–67.

9. Walker S, Mackay E, Barnett P, Sheridan Rains L, Leverton M, Dalton-Locke C, et al. Clinical and social factors associated with increased risk for involuntary psychiatric hospitalisation: a systematic review, meta-analysis, and narrative synthesis. Lancet Psychiatry. 2019;6(12):1039–53

10. Barnett P, Matthews H, Lloyd-Evans B, Mackay E, Pilling S, Johnson S. Compulsory community treatment to reduce readmission to hospital and increase engagement with community care in people with mental illness: a systematic review and meta-analysis. Lancet Psychiatry. 2018;5(12):1013–22.

11. NHS Digital Community Treatment Orders 2023-24/ Community Treatment Orders - NHS England Digital/ Accessed 26/02/25

12. Molyneaux E, Turner A, Candy B, Landau S, Johnson S, Lloyd-Evans B. Crisis-planning interventions for people with psychotic illness or bipolar disorder: systematic review and meta-analyses. BJPsych Open. 2019;5(4):e53.

13. de Jong MH, Kamperman AM, Oorschot M, Priebe S, Bramer W, van de Sande R….& Mulder C. al. Interventions to Reduce Compulsory Psychiatric Admissions: A Systematic Review and Meta-analysis. JAMA Psychiatry. 2016;73(7):657–64.

14. Henderson, C., Swanson, J. W., Szmukler, G., Thornicroft, G., & Zinkler, M. (2008). A typology of advance statements in mental health care. Psychiatric Services, 59(1), 63–71.

15. Lasalvia A, Patuzzo S, Braun E, Henderson C. Advance statements in mental healthcare: time to close the evidence to practice gap. Epidemiol Psychiatr Sci. 2023 Dec 6;32:e68. doi: 10.1017/S2045796023000835. PMID: 38053411; PMCID: PMC10803188.

16. Department of Health and Social Care. Modernising the Mental Health Act – final report from the independent review. London: DHSC; 2018.

17. Henderson C, Farrelly S, Moran P, Borschmann R, Thornicroft G, Birchwood M, et al. Joint crisis planning in mental health care: the challenge of implementation in randomized trials and in routine care. World Psychiatry. 2015;14(3):281–3.

18. Henderson C, Flood C, Leese M, Thornicroft G, Sutherby K, Szmukler G. Effect of joint crisis plans on use of compulsory treatment in psychiatry: single blind randomised controlled trial. Bmj. 2004;329(7458):136.

19. Thornicroft G, Farrelly S, Szmukler G, Birchwood M, Waheed W, Flach C, et al. Clinical outcomes of Joint Crisis Plans to reduce compulsory treatment for people with psychosis: a randomised controlled trial. Lancet. 2013;381(9878):1634–41.

20. Lay B, Kawohl W, Rössler W. Outcomes of a psycho-education and monitoring programme to prevent compulsory admission to psychiatric inpatient care: a randomised controlled trial. Psychol Med. 2018;48(5):849–60.

21. Lay B, Salize HJ, Dressing H, Rüsch N, Schönenberger T, Bühlmann M, et al. Preventing compulsory admission to psychiatric inpatient care through psycho-education and crisis focused monitoring. BMC Psychiatry. 2012;12:136.

22. Tinland A, Loubière S, Mougeot F, Jouet E, Pontier M, Baumstarck K, et al. Effect of Psychiatric Advance Directives Facilitated by Peer Workers on Compulsory Admission Among People With Mental Illness: A Randomized Clinical Trial. JAMA Psychiatry. 2022;79(8):752–9.

23. Kular, A., Birken, M., Wood, L., Parkinson, J., Bacarese-Hamilton, T., Blakley, L., … & Johnson, S. (2025). Exploring pathways to compulsory detention and ways to prevent repeat compulsory detentions in England; clinician perspectives. PLOS Mental Health, 2(6), e0000314.

24. Birken M, Kular A, Nyikavaranda P, Parkinson J, Mitchell L, Fraser KL, et al. Exploring pathways to compulsory detention in psychiatric hospital and ways to prevent repeat detentions; Service user perspectives. medRxiv. 2024:2024.06.04.24308425.

25. Johnson S, Birken M, Nyikavaranda P, Kular A, Gafoor R, Parkinson J, et al. A crisis planning and monitoring intervention to reduce compulsory hospital readmissions (FINCH study): protocol for a randomised controlled feasibility study. Pilot and Feasibility Studies. 2024;10(1):35.

26. Lean M, Fornells-Ambrojo M, Milton A, Lloyd-Evans B, Harrison-Stewart B, Yesufu-Udechuku A, Kendall T and Johnson S. Self-management interventions for people with severe mental illness: systematic review and meta-analysis. The British Journal of Psychiatry. 209; 214(5): 260–268.

27. Islam S, Appleton R, Hutchings-Hay C, Lloyd-Evans B, Johnson S. A systematic review of influences on implementation of supported self-management interventions for people with severe mental health problems in secondary mental health care settings. PLoS One. 2023;18(2):e0282157.

28. Lloyd-Evans B, Christoforou M, Osborn D, Ambler G, Marston L, Lamb D…& Johnson S Crisis resolution teams for people experiencing mental health crises: the CORE mixed-methods research programme including two RCTs. Programme Grants Appl Res 2019;7(1). 10.3310/pgfar07010

29. Craig P, Dieppe P, Macintyre S, Michie S, Nazareth I, Petticrew M. Developing and evaluating complex interventions: the new Medical Research Council guidance. Bmj. 2008;337:a1655.

30. Skivington K, Matthews L, Simpson SA, Craig P, Baird J, Blazeby JM, et al. A new framework for developing and evaluating complex interventions: update of Medical Research Council guidance. Bmj. 2021;374:n2061.

31. Hoffmann TC, Glasziou PP, Boutron I, Milne R, Perera R, Moher D, et al. Better reporting of interventions: template for intervention description and replication (TIDieR) checklist and guide. Bmj. 2014;348:g1687.

32. Thabane L, Ma J, Chu R, Cheng J, Ismaila A, Rios LP, et al. A tutorial on pilot studies: the what, why and how. BMC Med Res Methodol. 2010;10:1.

33. Eldridge SM, Lancaster GA, Campbell MJ, Thabane L, Hopewell S, Coleman CL, Bond CM. Defining Feasibility and Pilot Studies in Preparation for Randomised Controlled Trials: Development of a Conceptual Framework. PLoS One. 2016;11(3):e0150205.

34. NHS Digital (2018) Mental Health Act Statistics, Annual Figures England, 2017–18 https://files.digital.nhs.uk/34/B224B3/ment-heal-act-stat-eng-2017-18-summ-rep.pdf Accessed 16/12/22

35. Attkisson CC, Greenfield TK. Client Satisfaction Questionnaire-8 and Service Satisfaction Scale-30. The use of psychological testing for treatment planning and outcome assessment. Hillsdale, NJ, US: Lawrence Erlbaum Associates, Inc; 1994. p. 402–20.

36. Neil S, Kilbride M, Pitt L, Nothard S, Welford M, Sellwood W, Morrison A. The questionnaire about the process of recovery (QPR): A measurement tool developed in collaboration with service users. Psychosis. 2009;1:145–55.

37. Carpinello SE, Knight E, Markowitz F, Pease EA. The development of the mental health confidence scale: A measure of self-efficacy in individuals diagnosed with mental disorders2000. 236–43 p.

38. Keetharuth AD, Brazier J, Connell J, Bjorner JB, Carlton J, Taylor Buck E, et al. Recovering Quality of Life (ReQoL): a new generic self-reported outcome measure for use with people experiencing mental health difficulties. Br J Psychiatry. 2018;212(1):42–9.

39. Herdman M, Gudex C, Lloyd A, Janssen M, Kind P, Parkin D, et al. Development and preliminary testing of the new five-level version of EQ-5D (EQ-5D-5L). Qual Life Res. 2011;20(10):1727–36.

40. Overall JE, Gorham DR. The Brief Psychiatric Rating Scale. Psychological Reports. 1962;10:799–812.

41. Beecham J, Knapp M. Costing Psychiatric Interventions. Measuring mental health needs. 2001;2.

42. Developing and testing a strategy to prevent involuntary psychiatric hospital admissions. ISRCTN trial registry 10.1186/ISRCTN11627644

43. Sekhon M, Cartwright M, Francis JJ. Acceptability of healthcare interventions: an overview of reviews and development of a theoretical framework. BMC Health Services Research. 2017;17(1):88.

44. Lorenc, A., Rooshenas, L., Conefrey, C. et al. Non-COVID-19 UK clinical trials and the COVID-19 pandemic: impact, challenges and possible solutions. Trials 24, 424 (2023). 10.1186/s13063-023-07414-w

45. Gillard, S., Bremner, S., Patel, A., Goldsmith, L., Marks, J., Foster, R., … & Priebe, S. (2022). Peer support for discharge from inpatient mental health care versus care as usual in England (ENRICH): a parallel, two-group, individually randomised controlled trial. The Lancet Psychiatry, 9(2), 125–136.

46. Johnson, S., Lamb, D., Marston, L., Osborn, D., Mason, O., Henderson, C., … & Lloyd-Evans, B. (2018). Peer-supported self-management for people discharged from a mental health crisis team: a randomised controlled trial. The Lancet, 392(10145), 409–418.

47. Foye U, Lyons N, Shah P, Mitchell L, Machin K, Chipp B….& Simpson A. Understanding the barriers and facilitators to delivering peer support effectively in England: a qualitative interview study. BMC Psychiatry. 2025 May 12;25(1):480. doi: 10.1186/s12888-025-06850-z. PMID: 40355846; PMCID: PMC12070543.

